# Targeted Stimulation of the Sensory Afferents Improves Motoneuron Function in Humans with Spinal Muscular Atrophy

**DOI:** 10.1101/2024.02.14.24302709

**Authors:** G. Prat-Ortega, S. Ensel, S. Donadio, L. Borda, A. Boos, P. Yadav, N. Verma, J. Ho, S. Frazier-Kim, D.P. Fields, LE Fisher, DJ Weber, T. Duong, S. Weinstein, M. Eliasson, J Montes, K.S. Chen, P Clemens, P. Gerszten, G.Z. Mentis, E Pirondini, R. M. Friedlander, M Capogrosso

**Author notes:** Correspondence to: Marco Capogrosso, Robert Friedlander, Elvira Pirondini. co-first authors (G. Prat-Ortega, S. Ensel, S. Donadio). co-senior authors (E Pirondini, R. M. Friedlander, M Capogrosso).

## Abstract

Spinal Muscular Atrophy (SMA) is an inherited neurodegenerative disease causing motoneuron dysfunction, muscle weakness and early mortality^1,2^. Three therapies can slow disease progression enabling people to survive albeit with lingering motoneuron dysfunction and severe motor impairments^3,4^. Here we introduce a neurotechnological approach that improved spinal motoneuron function, muscle strength and walking in three adults with SMA. Starting from preclinical evidence showing that motoneuron dysfunction in SMA originates from the loss of excitatory inputs from primary afferents^5,6^, we hypothesized that augmentation of sensory neural activity with targeted electrical stimulation could compensate for this loss thereby improving motoneuron function. To test this hypothesis we implanted three adults with SMA with epidural electrodes over the lumbosacral spinal cord to stimulate the sensory axons of the legs^7,8^. We stimulated participants for 4 weeks 2 hours per day while they executed walking and strength tasks. Remarkably, our neurostimulation regime led to robust improvements in strength, walking and fatigue paralleled by reduced neuronal hyperexcitability, increased sensory inputs and higher motoneuron firing rates. Our data indicates that targeted neurostimulation can reverse degenerative processes of circuit dysfunction thus promoting disease modifying effects in a human neurodegenerative disease.

## MAIN

Disease onset and progression in chronic neurodegenerative diseases is commonly assumed to be driven by progressive neuronal cell death resulting in damage to neural circuitry impacting neurological function including cognitive, sensory and motor impairments^9–15^. In fact, in addition to neuronal cell death, synaptic circuit dysfunction is emerging as another determinant of deficits in Parkinson’s, Alzheimer’s, Huntington’s disease, ALS and Spinal Muscular Atrophy (SMA)^6,16–19^. Therefore, targeting neuronal death is not sufficient to reverse the disease phenotype. Instead attention must be brought to the design of therapies that also target neural circuit dysfunction. Given the existence of neuroprotective therapies for SMA^3,4,19–2^ and the basic neural circuits affected by SMA^6^, this disease is particularly suited to validate a combined disease modifying strategy targeting neuroprotection and neural circuitry modulation. SMA is an inherited spinal motor-circuit disorder caused by the homozygous loss of the SMN1 gene which results in ubiquitous deficits of SMN protein expression^1,23^. Lack of this protein leads to selective death of spinal motoneurons and widespread progressive muscle atrophy. The disease manifests with varying phenotypic severities that depend on the number of copies of the nearly identical SMN2 gene and range from premature infant death to normal life-span adults living with progressive motor deficits and disability^24^. In the last 7 years, three breakthrough therapeutics that upregulate the synthesis of SMN protein were approved, revolutionizing clinical care of SMA^3,4,20–22^. However, while these therapies effectively prevent the mortality of affected children and decelerate disease progression in adults, they prove ineffective in completely reversing motor deficits. Consequently, treated individuals exhibit severe, persistent motor impairments, leading to loss of function throughout development and into adulthood^,19,20^.

This situation likely stems from the fact that motor deficits in SMA are not exclusively caused by motoneuron death. Animal models of SMA show that severe motor deficits appear before widespread motoneuron death^6^, suggesting that spinal circuit dysfunction may play a role in disease onset and progression. Importantly, excitatory synaptic input to spinal motoneurons coming from primary proprioceptive afferents is significantly lower in SMA^5,6^. In turn, this excitatory synaptic deficiency causes SMA-affected motoneurons to undergo homeostatic compensation for loss of excitatory input that alter membrane ion-channels densities^27,28^. Unfortunately, these maladaptive homeostatic changes in membrane properties lead to hyperexcitable motoneurons that are paradoxically incapable of producing sustained action potential firing and in consequence normal muscle contraction^5,29^ (**Fig. 1a**).

**Figure 1:**
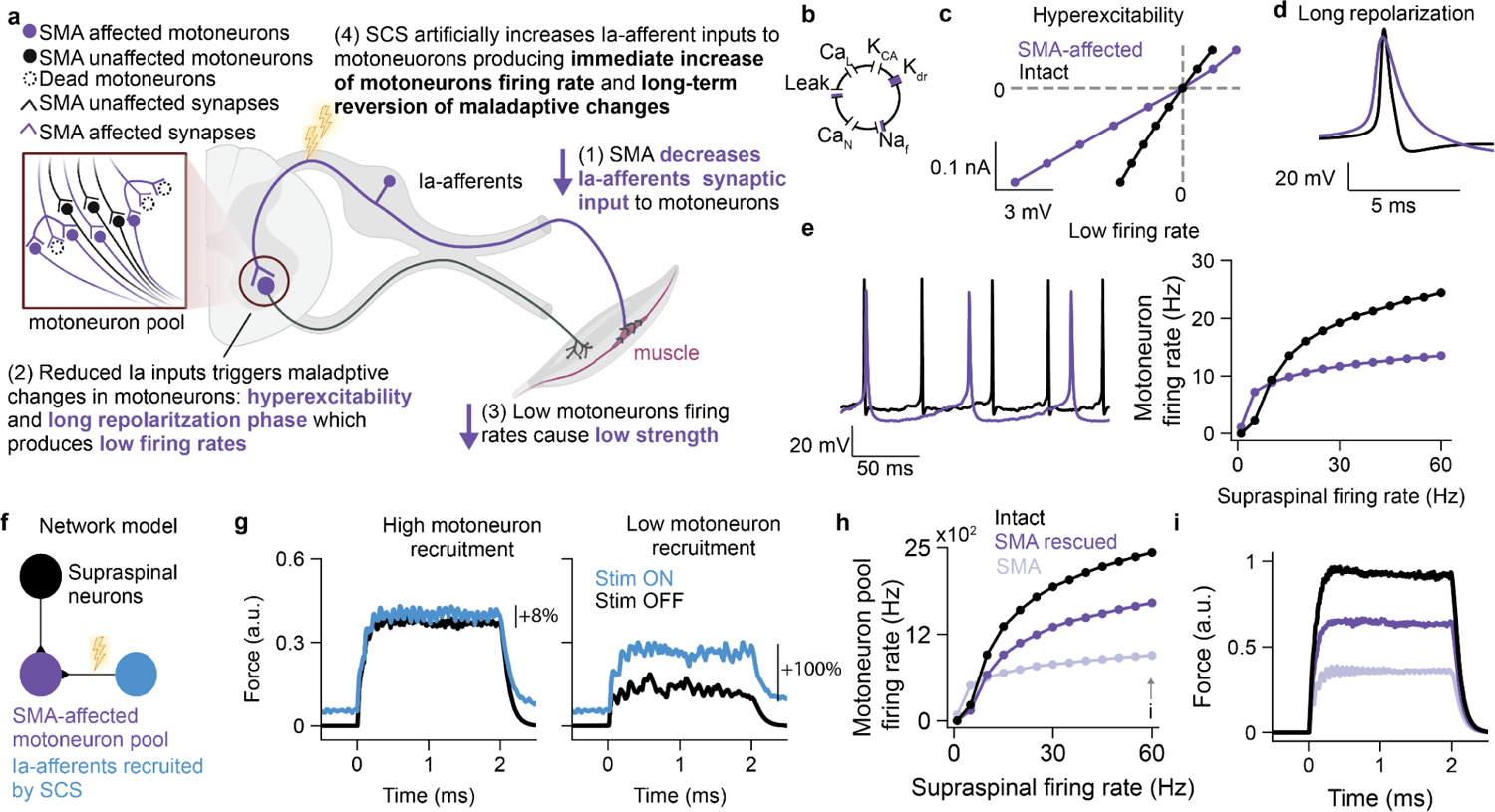
Hypothesis and theoretical framework **a,** Study hypothesis **b-d** A Hodgkin-Huxley model with a almost blocked delayed rectifier and partially blocked Sodium and Leak channels (**b**) reproduced the electrical properties of SMA-affected motoneurons^5^: Hyperexcitability characterized by a higher input resistance (**c**), Long repolarization phase after an action potential (**d**) and low firing rate (**e**). **f,** Network model schema: the SMA-affected motoneuron pool receives excitatory inputs from supraspinal neurons and Ia-afferents recruited by SCS. **g,** Assistive effect. Simulated force produced by an SMA-affected motoneuron pool with 30% motoneuron loss and 50% SMA-affected motoneurons with and without SCS. SCS amplitude: 11 Ia-afferents recruited, SCS frequency: 40Hz. The magnitude of the assistive effects depended on the recruitment of the motoneuron pool. For high motoneuron recruitment (supraspinal firing rate of 20 Hz) SCS produced only 8% increase in torque while for low motoneuron recruitment (supraspinal firing rate of 5 Hz) SCS produced 80% increase in torque. **h-i,**Therapeutic effect. Simulated firing rates (**h**) and forces (**i**) for a motoneuron pool with 30% of motoneuron loss. The biophysical model predicted a large increase of maximum firing rate and force when SMA-affected motoneurons are functionally rescued. However, the maximum force was still affected by motoneuron death.

Upregulating the activity of primary sensory afferents could compensate for the loss of excitatory input to motor neurons potentially triggering circuit dysfunction reversal^29^ (**Fig. 1a**). Studies in mice utilizing pharmacological agents suggest that while this hypothesis may be correct, the systemic delivery of non-specific drugs at doses required to upregulate inputs into spinal motoneurons can lead to systemic side effects making translation to humans difficult^29^. We therefore sought to overcome this safety concern by designing a human-safe intervention to achieve selective upregulation of spinal sensory afferents inputs by means of targeted electrical stimulation. Specifically, we hypothesized that we could use Spinal Cord Stimulation (SCS), a clinically approved neurostimulation therapy^30^, to selectively activate sensory afferents in the dorsal roots^8,31–34^ with electrical stimulation. Using SCS directed to vulnerable spinal segments, we aimed at demonstrating two hypothesis: 1) when turned ON, SCS can increase motoneuron firing rates, thereby immediately increasing strength and quality of movement, an assistive effect (i.e. an effect that is immediate, it assists movement but it disappears when SCS is turned OFF) and 2) over time, SCS would lead to the reversal of maladaptive homeostatic changes in the electrical properties of SMA-affected motoneurons thereby reversing motor deficits in humans providing a therapeutic effect (e.g. an effect that persists in time even when SCS is OFF).

### Predicted effects of SCS in a model of SMA-affected spinal motoneurons

We first aimed at assessing the theoretical validity of our hypothesis using computer simulations^31,34,35^. Specifically, we adapted a Hodgkin-Huxley model of spinal motoneurons’ membrane potential^36–38^ with data from mice with SMA to reproduce the biophysical signatures of motoneuron dysfunction in SMA: hyperexcitability, low firing rates and decreased synaptic inputs from Ia-afferents^5,6^. For this, we first adjusted the motoneuron model ion conductances to mimic the hyperexcitability of SMA-affected motoneurons characterized by a high input resistance (Methods, **Fig. 1b, c**). Second, we reduced the delayed rectifier potassium channel conductance which, paralleling experimental data, extended the action potential repolarization phase reducing motoneurons firing rates^5^. (Methods, **Fig. 1d, e**). Finally, to model the decrease in synaptic inputs from Ia-afferents, we reduced the strength of synapses from Ia-afferents^5^.

Experimental data show that not all surviving motoneurons are dysfunctional in SMA. Motorpools are composed of a mix of normal functioning units (SMA-unaffected) and dysfunctional units (SMA-affected). We used this property to simulate an SMA-affected motoneuron pool by reducing the number of motoneurons (mimicking motoneuron death) and mixing SMA-affected and SMA-unaffected motoneurons. This simulated motoneuron pool received inputs from supraspinal neurons to mimic volitional recruitment of motoneurons (**Fig. 1f**, Extended Data Fig. 1a). We then sent excitatory inputs from supraspinal neurons and found that the simulated motoneuron pool produced markedly lower forces than a fully-intact motoneuron population as expected in SMA. (**Extended Data Fig. 1b, c**).

We then used the model to check if SCS could yield beneficial motor assistance in SMA (hypothesis 1). For this, we modeled the effects of SCS on the network as a synchronous recruitment of Ia-afferents at the stimulation frequency as we demonstrated in the past^31,37–39^ (**Fig. 1f**). We found that when SCS was turned ON (40 Hz) motoneurons produced higher firing rates that translated into an increase in simulated forces compared to the SCS OFF condition (**Fig. 1g, Extended Data Fig. 1d, e**). However, these gains were higher at lower forces (+100%, i.e. when less motoneurons were recruited from supraspinal neurons) than at higher forces (+10% increase). In other words, if all motoneurons were volitionally recruited at their maximum firing rates, SCS could not further increase their firing rates (**Extended Data Fig. 1d, e**). Thus, our model predicts that SCS should be able to immediately increase motoneuron firing rates in SMA, however the magnitude of this assistive effect would depend on the effort exerted. This means that SCS would be more effective in daily living activities than in maximum voluntary muscle contraction.

Our second hypothesis was that if SCS is applied over time it would lead to the reversal of motoneuron dysfunction by means of increased sensory inputs. The rescue of motoneuron function would produce an increase in motoneuron firing capacity over time even without SCS, a therapeutic effect (**Fig. 1a**). Therefore, we studied the effect that rescuing some of the SMA-affected motoneurons (i.e. transforming SMA-affected into SMA-unaffected motoneurons) would have on simulated forces. We found that rescuing the function of even some of the SMA-affected motoneurons led to substantial improvements in strength (**Fig. 1h, i**, Extended Data Fig. 1b, c) that were much larger than the assistive effects of SCS. Thus our model predicted that SCS should be able to assist the production of movement when ON and that if it could rescue the function of some SMA-affected neurons it would lead to even higher improvements in strength (therapeutic effects).

### Quantitative assessment of the effects of SCS in humans with SMA

To empirically test our hypotheses we ran a first-in-human study with ambulatory adults living with SMA. We previously demonstrated that epidural SCS leads, when surgically placed dorsolaterally in the epidural space, can recruit sensory afferents in selected dorsal roots^8,31,33,40–42^. We leveraged this property to design a pilot study (NCT05430113) to assess the feasibility of using SCS to improve leg motor function in adults with SMA. We recruited three participants with lower-limb weakness, atrophy and impairments ranging from mild (SMA01) to severe (SMA02, **Extended Data Table 1, Extended Data Fig. 2a, d, g**) who received a temporary bilateral implant of two 8-contact linear epidural leads for 29 days (after which the leads were removed for a safety cautionary approach, **Fig. 2a**). We followed intraoperative mapping procedures previously described^8,40,43^ to position the electrodes above the spinal segments L1-S1 in order to engage the dorsal roots that innervate the leg muscles^8^ (**Fig. 2a**).

**Figure 2:**
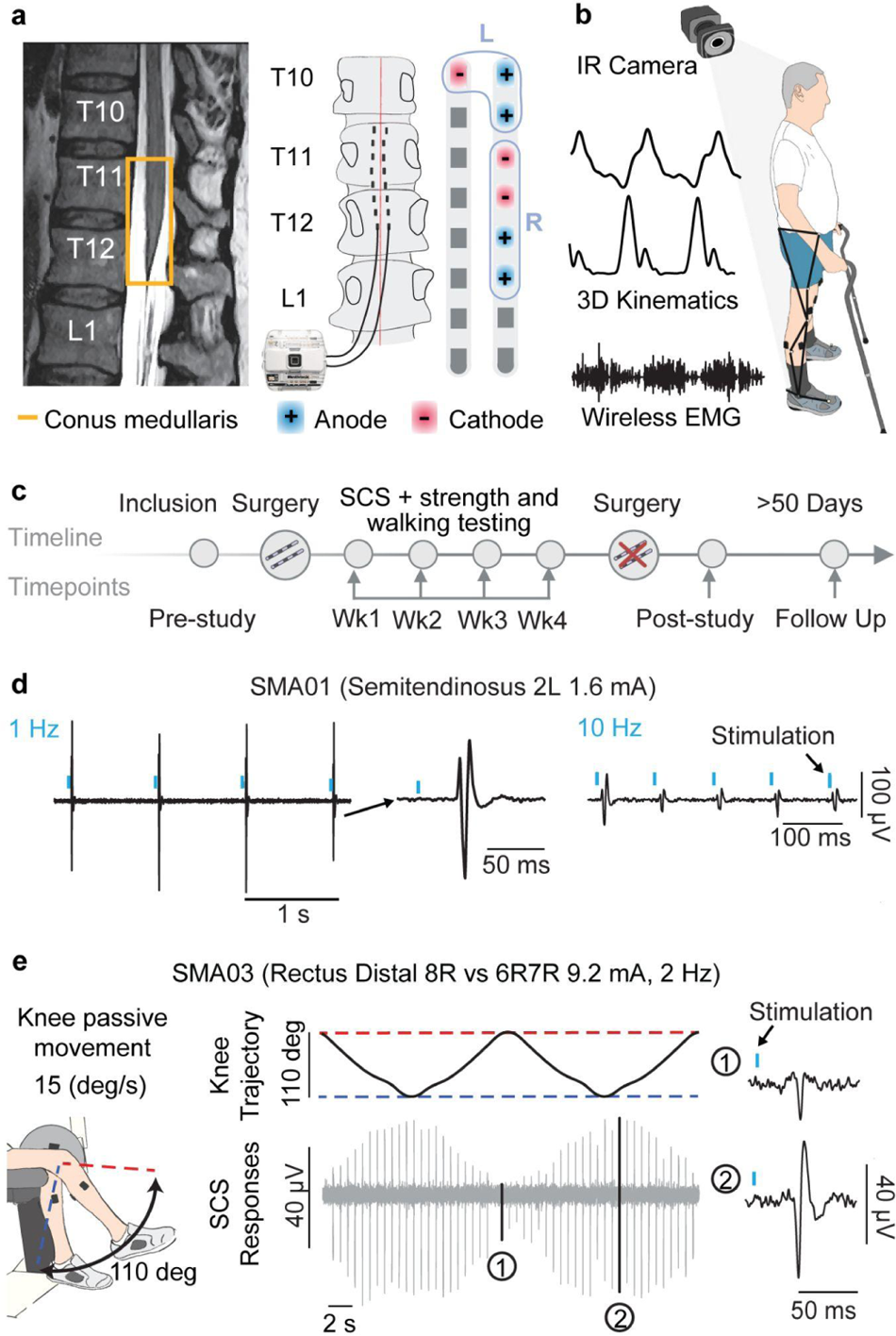
Study design and evidence for sensory recruitment with SCS **a**, Left: Example of T2 MRI (SMA02), the conus medullaris (highlighted in yellow) is located under the T12 vertebra. Center: The electrodes were bilaterally placed in the epidural space. For SMA02, they were placed between T11 and T12. In most of the experiments, we used a wireless stimulator connected to the leads with 8 contacts each. Right: Example of SCS parameters, where we used a program targeting the right (amplitude: 5 mA, frequency: 40 Hz and pulse width: 400 μs) and the left (5.5 mA, 40Hz and 400 μs) leg hip flexors and knee extensors muscles. **b**, We record 3D Kinematics and EMG Activity during Overground Walking **c**, Study Timeline. **d**, We sent SCS pulses at 1, and 10 Hz and present an example (SMA01) where we observed clear frequency dependent suppression (i.e. decrease of peak-to-peak responses as we increased frequency) which is a signature of sensory driven recruitment of MNs. **e**, Left: Example set up for passive knee joint movement (SMA03). The participants were secured to a robotic system that moved the knee joint passively within the reported range of motion. SCS parameters were configured to target a muscle that underwent stretching cycles during the joint movement. Center: joint angles during the 110-degree oscillation and the corresponding EMG activity of the rectus femoris. Spinal reflexes are clearly modulated by joint angles, indicating that SCS activates sensory afferents. Insets 1 and 2 show two spinal reflexes examples at maximum knee extension and flexion, respectively.

Using spinal cord MRIs, we observed that all three participants had the conus medullaris under the vertebra T12, a vertebra above what is normally observed (L1) thus leading to a different optimal implant location compared to protocols for spinal cord injury^8,43–45^ (**Fig. 2a**, Extended Data Fig. 2b, e, h). Following recovery, participants underwent 19 sessions of walking and knee strength testing during which we acquired a battery of kinematic, kinetic and electrophysiology assessments. All these experiments were geared to quantify the assistive and therapeutics effects of SCS as well as validating its mechanisms of action through imaging and electrophysiology (**Fig. 2b, c**). Assessments included 3D kinematics during walking, standard and high-density electromyography, single joint isometric torques, Transcranial Magnetic Stimulation, and functional imaging of the spinal cord^46,47^. Following recovery from surgery, we individualized SCS parameters to focus SCS towards the hip (the most affected muscles in SMA) and knee muscles following procedures that we previously described^7,8,40^ and used these configurations for the rest of the study (**Fig. 2a**, Extended Data Fig. 2c, f, i). Importantly, no-serious adverse events nor adverse events related to the use of SCS in humans with SMA were reported for the duration of the study (see supplementary information).

### SCS recruits spinal motoneurons via afferent reflex pathways in humans with SMA

Given the severe degeneration of spinal circuits in SMA, we sought to verify that SCS could indirectly engage surviving spinal motoneurons by directly stimulating sensory afferents in the dorsal roots rather than ventral axons (an important safety precautionary approach in motoneuron diseases^48,49^). To test for this we acquired evoked EMG waveforms by single SCS pulses delivered at different frequencies and found that EMG responses were suppressed at higher SCS frequencies (>10Hz) (**Fig. 2d**), a known electrophysiological signature of sensory-mediated recruitment of spinal motoneurons^33,40,50^. We corroborated this finding with a second experiment where we verified that evoked reflex responses by single pulses of SCS were dependent on joint angles, another known signature of sensory-driven recruitment of motoneurons^38^. Specifically, we measured peak-to-peak EMG amplitudes evoked by each SCS pulse during passive motion of the leg at controlled speed with our isokinetic robot and found that these responses had clear joint-angle dependencies demonstrating their physiological equivalence to H-reflexes (**Fig. 2e**, Extended Data Fig. 3).

### Neurostimulation of the spinal cord leads to large increases in muscle strength

We next evaluated the effects of SCS on leg muscle strength. We first quantified the assistive effects of SCS, and asked whether participants produced stronger forces acutely, by comparing SCS ON and SCS OFF in the same session. We inspected isometric torques produced during maximal voluntary contractions at the hip and knee joints (**Fig. 3**) with and without SCS in the same session. Importantly, we analyzed all joints where participants could reliably produce more than 4 Nm, below which the biomechanical noise of our system prevented consistent assessments. This was particularly relevant for SMA02 who could only produce detectable forces at the hip (**Extended Data Fig. 4**, Methods). As expected, all participants had significant weakness during maximal voluntary contraction (2 to 220 times weaker in knee extension and 1 to 100 times weaker in knee flexion than able bodied persons^51^). When SCS was turned ON (at motor threshold, 40Hz), we observed facilitation in all joints in the participants that we tested in SMA01 and SMA03 but not all function (i.e. extension or flexion). Specifically, we measured immediate gain in strengths (up to +20%) at the hip and knee joints in both subjects (**Fig. 3b-d**). Interestingly, in SMA01 we observed consistent increases in knee extension strength when the participant started to fatigue (**Extended Data Fig. 4b**). Participants reported that movements felt easier when SCS was ON. Since they could always feel that SCS was active^40^ we could not execute a blinded control, however they were not able to distinguish among different SCS parameters. Therefore to rule out placebo effects we executed sham controls by changing SCS parameters without subject knowledge and demonstrated that the effects vanished if non-optimal SCS parameters were used in agreement with previous literature^40^ (**Extended Data Fig. 5c**). The size and consistency across joints of these effects confirm our simulations that assistive effects were present in the muscles that we targeted with SCS (hip and knee) even if participants were producing their maximum strength.

**Figure 3:**
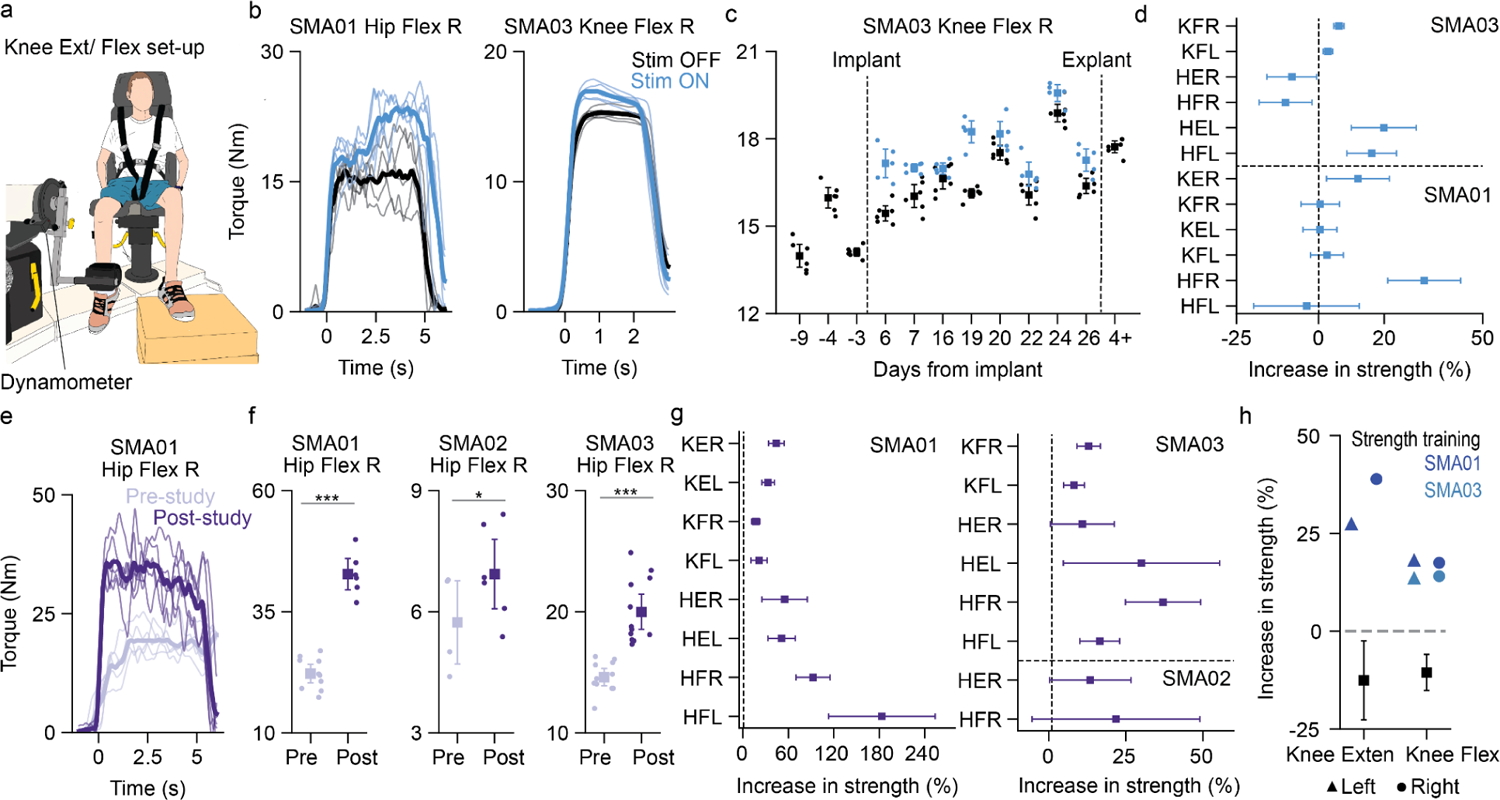
SCS improves muscle strength **a,** Experimental set up to measure isometric torques for knee extension and flexion. **b,** Examples of isometric torque traces during maximum voluntary contraction with and without SCS, light lines correspond to single trials while thick lines are the means across trials. **c,** Maximum torque produced by SMA03 during right knee flexion across days with and without SCS. **d,** Mean assistive increase in strength (SCS ON vs OFF, same day) for all muscles and participants that we could reliably measure (torques above 4 Nm). **e,** Example (SMA01) of isometric torque traces during maximum voluntary contraction pre and post study without SCS. **f,** Maximum torque produced for hip flexion right for all participants pre-study (pre-implant and week 1 trials) and post-study (week 4 and post-explant trials). One sided test using bootstrap with N=10,000 samples: *p<0.05 and ***p<0.001. **g,** Therapeutic increases in strength for all muscles and participants that we could reliably measure (torques above 4 Nm). **h,** Mean increase in strength for knee extension and flexion across participants after 3 months of aerobic and strength exercise^88^ (black) compared to the increase in strength for SMA01 and SMA03 at the same joints. For all panels, dots correspond to single trials and squares to the mean across trials. All error bars indicate the 95% confidence interval, computed with bootstrap N=10,000. Abbreviations: K, knee; H, hip; F, flexion; E, extension; R, right; L, left.

While executing these tests we noticed marked changes in torques even without SCS from a week to another. Specifically, we found large improvements pre vs post-study in almost all leg muscles (**Fig. 3g, h**, Extended Data Fig. 4), and particularly at the hip flexors (**Fig. 3e, f**) in all participants (up to +180% in hip flexion for SMA01). This is particularly remarkable in SMA02 who had severe muscle degeneration. Importantly, these therapeutic improvements largely exceed the extent of assistive effects and led over time to substantial motor benefits. For example, with assistive effects mounting on top of the newly acquired strength, SMA01 could stand up from a kneeling position when SCS was ON by week 4, and SMA03 could stand up from hinging on a desk by week 4, tasks that both participants were not able to do prior to the beginning of the study. To control for the effect of exercise without SCS we compared our strength results with data from the only randomized clinical trial performed on the same patient population (n=11 subjects) that underwent comparable intensity of strength and locomotion exercises^52,53^ (3120 min in control study vs 1400 min in our study) for which we had consistent measurements of knee strength with hand held dynamometry (HHD, **Fig. 3h**). Controls performing exercise only did not improve strength even after 3 months of regular exercise. In fact, the mean knee strength slightly decreased (extension: −12.5%, (−22.6,−2.5) 95% CI, flexion: −10.5%, (−15.1, −6.0) 95% CI). Our data falls entirely outside the confidence interval of this control dataset suggesting that observed changes in strength cannot be explained by exercise only (**Fig. 3g**). In summary, we found assistive effects on strength when SCS was turned ON that were paralleled by large changes in strength without stimulation that appeared over four weeks of SCS therapy.

### Neurostimulation of the spinal cord leads to improvements in fatigue and walking

We then tested whether the changes in strength were paralleled by similar improvements in functional locomotion as well as fatigue, a critical issue in SMA^54^. To assess the quality of gait we measured step height, step length, stride velocity (i.e. parameters associated with fatigue-related gait changes in SMA^55,55^) as well as range of motion at the hip and knee joints during overground walking using a 3D kinematic system (Vicon, USA). Following the trend observed in strength, when we turned SCS ON, all participants significantly changed their gait patterns as shown by changes in gait quality variables suggesting an important facilitatory effect on critical joint function for walking. SMA01 improved step length and stride velocity (**Extended Data Fig. 5b**), SMA02 improved in step height, step length and stride velocity, all participants increased their hip and knee joint range of motion (**Fig. 4a, b**, Extended Data Fig. 5a). These increases in range of motion were a direct consequence of an increase in the activation of the muscles involved (**Extended Data Fig. 6**). To further demonstrate this, we asked SMA01 who could sustain longer walking periods to exacerbate his hip flexion during weight-supported treadmill walking (**Extended Data Fig. 5f**). We then turned SCS ON and OFF during walking and observed an immediate large increase in step height that was directly linked to SCS being ON or OFF. To control for placebo effects we again performed control assessments by delivering sub-optimal SCS parameters and demonstrated that gait quality benefits were SCS parameter dependent (**Extended Data Fig. 5d**). Overall these results demonstrate that even small assistive effects on strength led to functional changes in locomotion.

**Figure 4:**
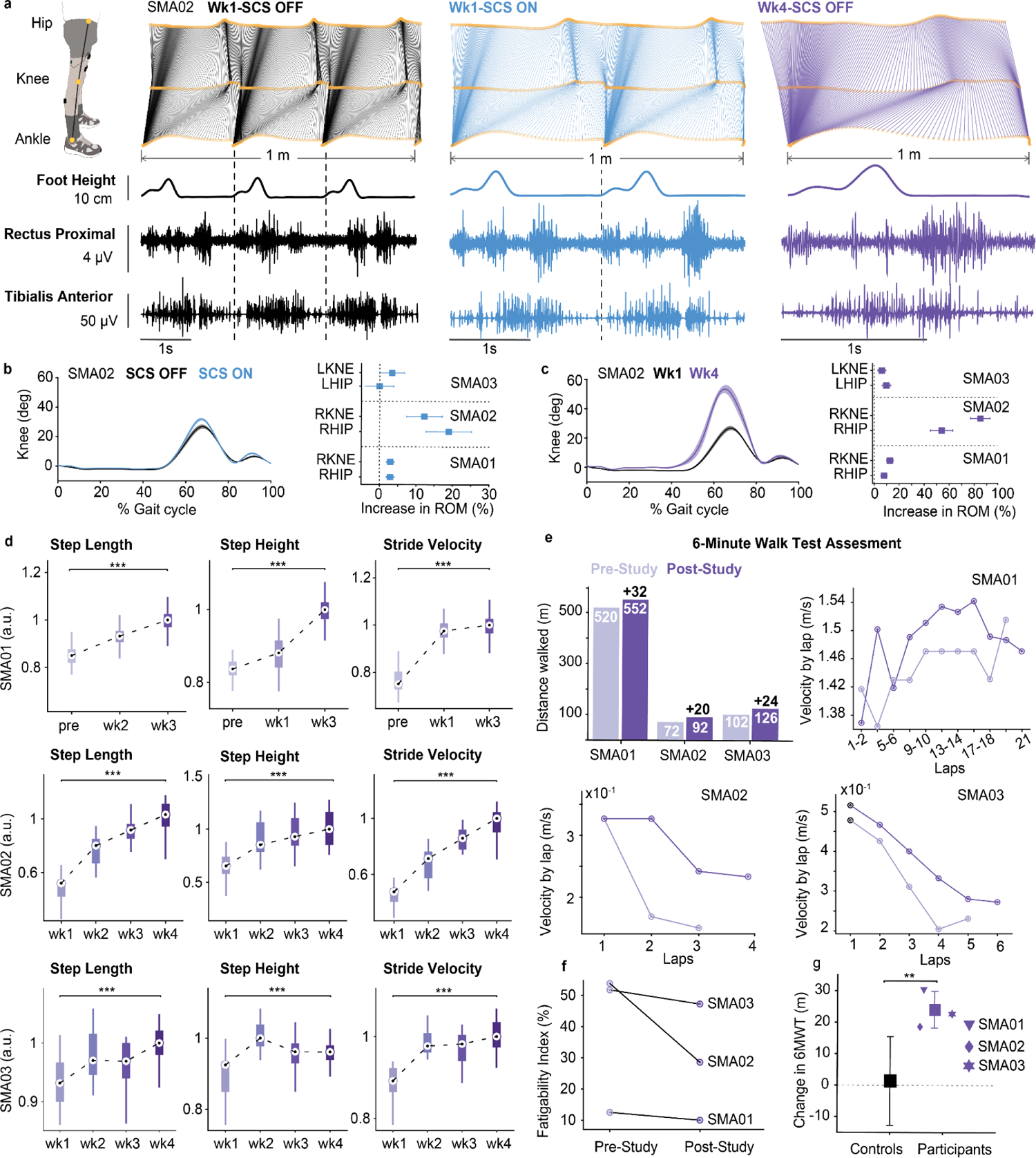
SCS improves gait quality **a**, Stick diagrams showing an example (SMA02) of the walking pattern across a fixed distance under varying conditions: SCS OFF (black), SCS ON (light blue) in week 1, and SCS OFF (purple) in week 4. Foot height and electromyography (EMG) activity for the Rectus Femoris proximal and Tibialis Anterior during the same distance walked. In this example, SMA02 exhibited a reduction in required strides to advance 1 m from three without SCS to two with SCS in week 1, and a single stride with SCS off at the end of study. **b**, Left: percentage assistive increase in hip and knee Range of Motion for all participants. Right: Mean and 95% CI of knee range of motion throughout the gait cycle with and without SCS in week 1 (assistive effect). **c**, Same as b but comparing week 1 and week 4 without SCS (therapeutic effect). **d**, Gait quality variables (step length, step height, and stride velocity) across weeks without SCS. Box plots, depicting the median and the 25th and 75th percentiles, the whiskers span the minimum and maximum data points, excluding outliers. **e**, Endurance and Fatigability analysis of the 6-Minute Walk Test (6MWT). Left: pre- and post-study 6MWT performances for all participants. Right: Velocity of each lap (∼20m) walked in the 6MWT for all participants. For SMA01, each data point represents the mean velocity computed between two consecutive laps, while for SMA02 and SMA03 each data point is a single lap. The decrease in velocity as a function of laps completed was smaller in the post-study phase, indicating a decrease in fatigability. **f**, Fatigability index pre and post-study computed as the percentage of change between the velocity of the first and last lap. **g**, Changes in the 6-Minute Walk Test (6MWT) pre vs post-study for all participants were outside the 95% confidence interval (−12.6,15.4) of a control group which underwent a three-month training protocol. Statistical significance was assessed with two-tail bootstrapping (N=10,000): p<0.05 (*), p<0.01 (**), p<0.001(***).

We then examined the therapeutic effects of SCS by looking at how these gait quality variables changed over time. All participants largely and significantly increased all gait quality variables over the 4 weeks of the study (**Fig. 4d**). Changes in gait quality were so prominent that SMA02 who had severe muscle degeneration and prior to the trial did not flex the knee at all, completely changed his gait pattern being able to fully flex the knee (+85% ROM at the knee and + 54% ROM at the Hip, **Fig. 4c**). Again these improvements in range of motion were paralleled by an increase in muscle activation of the muscles involved (**Extended Data Fig. 6**).

We then measured the impact of therapeutic improvements on fatigue and endurance using the standardized clinical measure of the 6 Minute Walk Test (6MWT)^54^. Remarkably, all our participants improved by at least 20m in the 6MWT^53^ (SMA01: +6.1%, SMA02: +27.7%, SMA03: +23.5% **Fig. 4e**, **Extended Data Table 1**). Since changes in 6MWT may reflect improvement in fatigability we examined lap-by-lap gait velocity, and compared the difference in the first and the last lap of the 6MWT to assess patterns of fatigability^55,56^ and found that all three participants had significant reduction in fatigability (**Fig. 4f**, −20% for SMA01, −47% SMA02 and −9% SMA03). This is markedly different from previously reported values that show a dissociation between improvement of the 6MWT and fatigue, whereas in our study all participants improved their 6MWT score and their fatigue score showing an intrinsic change in their motor phenotype^57^. Changes in SMA01 were so large that he noticed it in its life outside the lab, being now able to walk back from patient housing to the lab without fatiguing. We compared these results with the control dataset in people with SMA executing 3 months of regular exercise without SCS^52^ (see methods, **Fig. 4g**). Control individuals with ambulatory SMA undergoing comparable exercise regime showed a mean improvement of 1.4m over 3 months. Importantly, data from our study is outside the confidence interval of the controls’ distribution, demonstrating that the changes that we observed cannot be explained by existing data in the same patient population undergoing only physical exercise. In summary, we have shown that SCS led to large changes in fatigue and gait quality impacting functional motor behavior.

### Neurostimulation of the spinal cord improves spinal motoneuron function

Next we studied the mechanisms responsible for these large effect sizes to confirm our leading hypothesis that SCS reversed motoneuron dysfunction produced by SMA (**Fig. 1a**).

To directly assess this hypothesis, we used High-Density Electromyography^58^ (HDEMG, see methods) during isometric maximal voluntary contractions and extracted single motoneuron discharges for the knee extensors and flexors. First, we checked whether motoneurons of our SMA participants showed signatures of SMA-dysfunction: i.e. reduced peak firing rates. Motoneurons from able-bodied humans regularly reach peak values above 100 Hz^59–61^ at maximal contraction in the initial bursts. Instead, the mean peak firing across our participants was 50.6 Hz (95% CI 46.9, 54.3) confirming the presence of a substantial reduction in maximal repetitive firing of human motoneurons in SMA. We then assessed the assistive effects of SCS on motor units firing rates. We found that when SCS was turned ON some of the detected motor units increased their firing rates which improved mean population firing (**Fig. 5a**). This correlates to the immediate changes in torques (**Fig. 3d**) that we observed in experiments and predicted with simulations (**Fig. 1g**). We then looked at changes in motor units firing patterns that might have occured over time. When we compared motor units firing rates pre-vs post-study, we noticed that most of the post-study motor units firing dynamics was consistent with pre-study firing dynamics (**Fig. 5b**). However, new post-study motor units emerged that appeared to have a markedly different behavior. These units showed significantly higher peak firing rates (70.9 Hz., 95% CI 67.4, 74.4) at the onset of the force production that decayed over time rather than constant firing (**Fig. 5b, Extended Data** Fig. 7c). The distribution of peak firing rates for these units was markedly different from any other unit recorded pre-study (**Fig. 5b, Extended Data** Fig. 7b) therefore we labeled these units as *rescued units* since they demonstrated the acquired ability to fire at significantly higher rates much closer to able-bodied reference values.^59^ We found rescued units with remarkably similar firing dynamics in all three participants despite their large differences in disease progression suggesting that SCS promoted the reversal of motoneuron dysfunction in at least some of the SMA-affected surviving motoneurons in humans. These changes in motoneuron firing dynamics appeared over time and help explain the therapeutic changes in force that we observed in our data. If new high-firing motoneurons emerged, then this fact should be reflected into indirect measures of neural activity such as BOLD signal in functional MRI (fMRI) in the spinal cord. To test this, we designed fMRI sequences measuring BOLD signal changes in the lumbosacral spinal cord while participants volitionally flexed and extended their knee (**Fig. 5c-e, Extended Data** Fig. 8). The fMRI images produced clear BOLD signal activation that matched the lumbosacral position of motoneuron controlling the knee muscles (**Fig. 5c**). Remarkably, BOLD signal significantly increased (i.e., the z-score of active voxels) post-study compared to pre-study in all three participants indicating a measurable increase of neural activity in the spinal cord (**Fig. 5d, e**). In summary, we identified the emergence of motor units with firing capacities much closer to reference values for able-bodied humans paralleled by a significant increase in spinal BOLD fMRI signal during knee joint flexion and extension.

**Figure 5:**
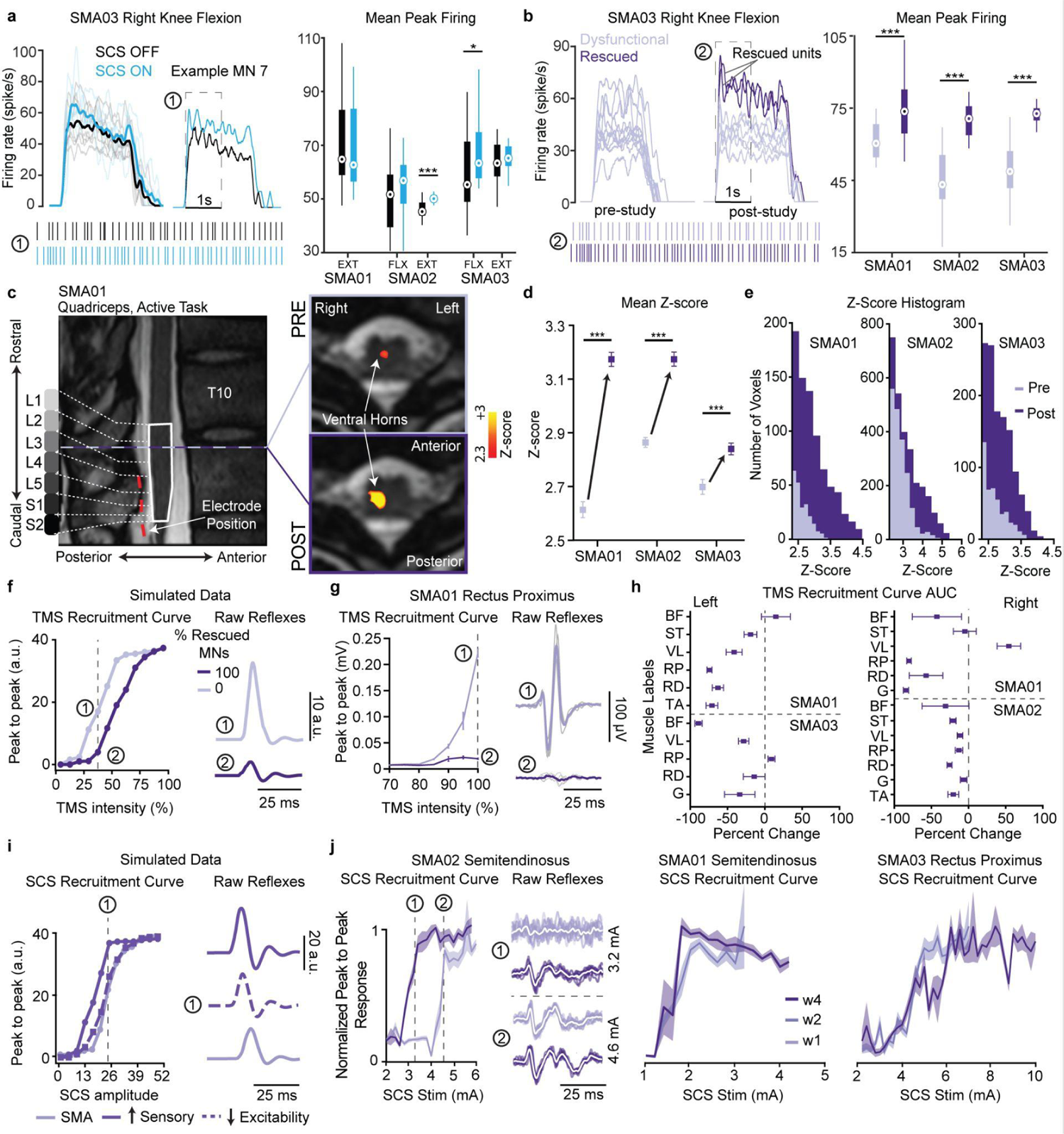
Electrophysiological evidence of improvements in motoneuron function. **a**, Left: traces a of single motoneuron firing rate during a maximum voluntary contraction in isometric condition with and without SCS. Raster plots show the spike times of two example motoneurons during the first second. Quantification of the mean peak firing rates across all isometric trials for SCS ON and OFF **b,** Left: traces of single unit MN firing rate for pre vs post-study. Post-study we found rescued motoneurons characterized by a higher peak firing rate. Right: Peak firing rate of pre-study vs rescued motoneurons **c,** Spinal segments are reported in the T2 anatomical image of SMA01. Thresholded activity patterns resulting from the generalized linear model are co-registered to the anatomical image from single pre and post scan sessions with maps presented uncorrected (Z > 2.3, p < 0.01). **d-e,** Quantification of the mean z-score (**d**) and histogram z-scores (**e**) of the L1-S2 spinal segments thresholded activity patterns maps. **f,** Simulated peak to peak response to a TMS pulse. The inset shows examples of simulated raw reflexes. The biophysical model predicted a decreased response to TMS stimulation with the number of rescued motoneurons. **g**, Example of TMS recruitment curve (SMA01, right rectus proximus). The inset block shows the raw waveforms at 100% intensity pre-vs post-study. **h,** Quantification of percent change in area under the curve of peak-to-peak TMS responses pre-vs post-study across all intensities tested. Abbreviations: ST, Semitendinosus; BF, Bicep Femoris; VL, Vastus Lateralis; RP, Rectus Femoris Proximal; RD, Rectus Femoris Distal; G, Gastrocnemius; TA, Tibialis Anterior. **i,** Biophysical model SCS recruitment curves for SMA-affected MN pools. The biophysical model predicts that the reflexes should increase as the number of rescued Ia-afferents increases but this effect can be compensated by the number of rescued motoneurons. Light purple: 0% rescued motoneuron and sensory afferents. Solid purple: 50% rescued motoneurons and Ia-afferents. Dash purple: 70% rescued motoneurons and 25% rescued sensory afferents. **j,** Examples of normalized recruitment curves for all participants. The inset block shows raw waveforms at two different amplitudes normalized for SMA02. In all box plots, the central circle indicates the median while the bottom and top edges of the box indicate the 25th and 75th percentiles, respectively. The whiskers extend to the minimum and maximum data points, not including outliers. Statistical differences were assessed with bootstrap (N=10,000). *, **, and *** denote significant differences with p-values of p < 0.05, p < 0.01, and p < 0.001, respectively.

### Neurostimulation of spinal cord alters motoneuron’s excitability and increases monosynaptic sensory synaptic strength

We hypothesized that SMA-affected motoneurons have reduced synaptic inputs from Ia-afferents, and in consequence increased excitability and low firing rates (**Fig. 1a**). Therefore, if it’s true that SCS reversed motoneuron dysfunction in SMA, then the increase in motoneuron firing rates that we observed should be paralleled by a decrease in motoneuron excitability and an increase in sensory synaptic inputs. Therefore we aimed at verifying these two remaining properties with electrophysiology. We first tested for motoneuron excitability using Transcranial Magnetic Stimulation (TMS). Indeed, in humans cortical neurons which have monosynaptic connections to spinal motoneurons are not affected by SMA^62^. Since SCS does not stimulate the cortico-spinal tract^33^, this pathway is not affected by our intervention and constitutes an independent pathway to test motoneuron excitability. In computer simulations, the rescue of motoneuron function led to significant decrease in simulated peak-to-peak Motor Evoked Potentials (MEPs) produced by single pulses of TMS (**Fig. 5f**). We then stimulated the leg-area of motor cortex pre- and post-study with single TMS pulses in all three participants. As expected we observed a significant, and large decrease in MEPs peak-to-peak amplitudes in all three participants (**Fig. 5h**) that rebounded rapidly at 6 weeks post-explant follow up (**Extended data Fig. 9a**). At the same time detected motor units numbers did not change (**Extended Data** Fig. 7d) which suggests that during the study the intrinsic excitability of spinal motoneurons was decreased, making it harder for TMS pulses to induce MEPs.

We then evaluated changes in sensory-to-motoneuron synaptic strength. We analyzed the sensory reflexes elicited by single pulses of SCS in leg muscles in the first and last week of implant. We found that reflexes were either unchanged or increased in all three participants (**Fig. 5i, j, Extended Data** Fig. 9). Consistent with simulations of sensory reflexes (**Fig. 5i**), this result implies an increase in sensory driven excitation to spinal motoneurons concurrent to the decrease in motoneuron excitability measured above (**Fig. 1a**). Reflexes should either remain stable or increase in consequence of the balancing effects of increased excitatory inputs and decreased excitability. Together these findings constitute first proof that electrical stimulation altered sensory-to-motoneuron synaptic circuit strength which in turn improved neural function by normalizing motoneuron membrane excitability in humans with SMA.

## DISCUSSION

In this study we demonstrated that electrical stimulation of the sensory afferents alleviates motor deficits in humans with SMA and, importantly, improves spinal motoneuron function in an otherwise progressive neurodegenerative disease.

We built our study around the hypothesis that motor deficits in SMA were the result of two independent phenomena, motoneuron death and spinal circuit dysfunction^5,6,29^ and aimed at defining a therapy that could reverse circuit dysfunction. Our combined behavioral, electrophysiological and imaging analysis shows that continued stimulation of the Ia-afferents altered motoneuron properties at a membrane level resulting in significant increases in spinal motoneuron activity that paralleled large and rapid changes in muscle strength and gait quality variables. Our finding is consistent with the hypothesis that people living with SMA have a set of surviving motoneurons, some or most of which have reduced firing capabilities that can be rescued by targeted neurostimulation. This suggests that targeted electrical stimulation can alleviate motor deficits by reversing the maladaptive homeostatic processes^27^ triggered by spinal circuit dysfunction. Evidence of improved motoneuron function was consistent across all three participants, irrespectively of their state of disease progression, age and of them being or not under SMN inducing therapies (SMA01 was not taking any therapy up until the end of our study). This shows that the results that we obtained were independent from the neuroprotective interventions and reinforce the hypothesis that motoneuron death and dysfunction in SMA are two independent consequences of the disease^5^. Actually, the different nature between SCS and existing therapies (SCS directly tackles the SMA circuit dysfunction while current therapies induce SMN production) suggests that a synergistic intervention that combines both may lead to accelerated recovery and real impact in daily life for people living with SMA.

The strong motor improvements that we observed occurred in a clinical population that, similarly to other motoneuron diseases, is well known for not responding to physical exercise^52,53,63^. In fact, statistical analysis against data from the only randomized clinical trial in adults with SMA exploring the effect of exercise demonstrates that our results cannot be explained by physical exercise only. Interestingly, SMA02 participated in both studies, allowing us to compare his specific results across studies. In the exercise only study his 6MWT did not change (−6m). Instead, in our study, he improved by 20m (+27.7%). This isn’t to say that exercise did not contribute in any way to the observed functional effects. However, our data clearly shows that electrical stimulation led to large quantitative changes in spinal motoneuron function in people with a degenerative motoneuron disorder: a disease-modifying effect. To provide context for our functional gains, we obtained a median of +24m, minimum +20m and maximum +34m changes in 6MWT scores in 4 weeks. Recent reports on the efficacy of the clinically approved SMN inducing drug Nusinersen in ambulatory adults^64^ show that people gain a median of 20 minimum of 9m and maximum of 48m in the 6MWT after 15 months. This means that our data is comparable with efficacy of approved clinical treatments but obtained in only 4 weeks. Moreover, we did not observe evidence of saturation or ceiling in improvements of both strength (**Fig. 3c**) and kinematics (**Fig. 4d**) suggesting that a longer treatment may yield even larger effects. Finally, follow up at 6 weeks of TMS and 6MWT data suggest that SCS must be administered to retain the improvements in motoneuron function.

In summary, our results provide insights into the disease mechanisms of SMA that lead to circuit and motoneuron dysfunction in humans. Importantly, we leveraged the identification of these mechanisms to design a clinically relevant intervention that manipulated the maladaptive processes induced by SMA to reverse neural dysfunction and improved function at a cellular, circuit and behavioral level. SCS is being studied by us and others as an assistive neuroprosthetic tool to improve movement after spinal cord injury^8,44,45^, stroke^40^ and even neurodegenerative diseases^65–67^. However this is the first time that a neurostimulation therapy was not engineered to assist movement, but to reverse degenerative circuit processes and effectively rescue motoneuron function in humans, a disease-modifying effect. Whether the size and consistency of these beneficial effects will be sustained over long-term use of neurostimulation in humans is unknown and should be explored in a follow up, controlled clinical trial. In conclusion, we demonstrated that targeted neurostimulation, alone or in combination with existing therapeutics, harbors the potential to halt disease progression, thereby altering the course and clinical management of neurodegeneration.

## Data Availability

All data produced in the present study are available upon reasonable request to the authors

## ACKNOWLEDGMENT

This study was supported by an exploratory research grant from F. Hoffmann–La Roche to MC and RMF. We wish to thank Sydney Bader for the support as clinical coordinator for this work. We wish to thank the participants of this study for being pioneers in neurostimulation research and for the great insight into the real needs of people living with SMA. We also wish to thank Dr. Livio Pellizzoni at Columbia University for the engaging and useful conversations on the mechanisms of SMA. Finally we wish to thank the SMA Foundation and specifically Loren Eng for her irreplaceable advice, support and encouragement.

## AUTHOR CONTRIBUTIONS

MC, RMF and ME conceived the study; GPO, SE and SD performed and designed all experiments and analyzed the data; LB, NV performed single motoneuron experiments, GPO, SE, SD and LB created figures; LB, PY, NV and DW analyzed single motoneuron data; AB performed all clinical evaluations under the supervision of TD, JM and PC; SFK analyze physical activity data; DF and PG implanted all participants and designed implant procedures with MC and LEF; ME, SW, KSC, MC and TD designed the clinical study; PC directed participants assessments and eligibility; JH performed and analyzed TMS data; SE and EP conceived and performed all fMRI experiments and analysis; JM provided the control dataset and supported the interpretation of clinical outcomes; GPO and MC conceived and implemented the computational model; MC, EP, RMF and GZM directed the scientific interpretation of data; RMF, EP and MC directed all experimental activities; RMF and MC secured funding for the study; GPO, SE, SD, EP, MC and RMF wrote the paper and all authors contributed to its editing.

## METHODS

More information about the clinical trial, the surgical procedures, EMG Acquisition, the SCS stimulators and MRI sequences can be found in Supplementary Information.

### Trial and participant information

All experimental protocols were approved by the University of Pittsburgh Institutional Review Board (IRB) (protocol STUDY21080158) under an abbreviated investigational device exemption. The study protocol is publicly available on ClinicalTrials.gov (NCT05430113). Three male individuals actively took part in the study, participating in every experiment. When participants had specific limitations, we adjusted certain procedures accordingly, clearly indicating such cases. Prior to their involvement, participants underwent an informed consent process, in accordance with the procedure approved by the IRB. Participants received compensation for each day of the trial, as well as for their travel and lodging expenses during the study period.

### Inclusion criteria

Individuals aged 16 to 65, diagnosed with 5q-autosomal recessive Spinal Muscular Atrophy (SMA) confirmed through genetic testing for a deletion in the SMN1 gene (5q12.2-q13.3), were eligible for participation if the disease manifested after 18 months of age and after the acquisition of ambulation (Type 3 or Type 4 SMA). All participants were required to be capable of standing independently for at least 3 seconds and have a pre-study Revised Hammersmith Scale (RHS) score equal to or lower than 65. Before enrollment, individuals underwent a medical evaluation for screening. Those with severe comorbidities, implanted medical devices preventing magnetic resonance imaging, claustrophobia, or those who were pregnant or breastfeeding were excluded from the study. Throughout the study period, participants were not allowed to take any anti-spasticity, antiepileptic, or anticoagulation medications.

### Study design and data reported

The objective of this exploratory clinical trial is to obtain preliminary evidence of safety and efficacy of SCS as a potential treatment to enhance motor function in individuals diagnosed with Type 3 or 4 spinal muscular atrophy (SMA). The study is structured as a single-center, open-label, non-randomized trial. We expect to enroll up to six subjects exhibiting quantificable motor deficits in the legs but capable of independent standing. Given the pilot nature of the study, SCS leads are implanted for a maximum of 29 days to minimize safety risks, after which the electrodes are removed. The primary and secondary outcomes are designed to primarily assess safety and obtain initial clinical and scientific evidence regarding both the assistive and therapeutic effects of SCS (See Supplementary Information).

Following screening and establishment of pre-study baselines, participants undergo implantation of percutaneous, bilateral, linear spinal leads near the lumbar spinal cord. Scientific sessions are conducted five times per week, lasting 4 hours each, totaling 19 sessions, starting from day 4 post-implant. Tasks and measurements in the initial two weeks focus on identifying optimal SCS configurations, which are then maintained for the remaining sessions. The primary outcomes aim to assess safety by monitoring adverse events related to SCS use. Participants rate the “discomfort/pain” on a scale of 1 to 10 for each SCS configuration to ensure that SCS intensities required for motor function improvement remain within a range of non-painful sensations. Additionally, the primary outcomes evaluate the effects of SCS on specific motor control variables. We quantify assistive and therapeutic improvements in strength by measuring isometric torque with and without SCS regularly during the trial. We rate motor deficits by assessing the Revised Hammersmith Scale (RHS), Hammersmith Functional Motor Scale Expanded (HFMSE), and the 6-minute walk test pre and post-study. We then evaluated function by measuring 3D kinematics during overground walking with and without SCS. Secondary outcomes aim to acquire scientific evidence of mechanisms that may contribute to improved motor performance. A battery of imaging and electrophysiology tests assess early signs of neural plasticity in the central nervous system associated with measured effects on motor control. A comprehensive description of primary and secondary outcomes can be found on <u>ClinicalTrials.gov (NCT05430113)</u>. This manuscript reports results from the first three participants.

### Participant information

In this work, we report the results from the first three individuals participating in our trial. All participants were diagnosed with SMA type 3 and they were able to stand and walk (with the assistance of walking aids for SMA02 and SMA03). SMA01 (age range 21-25 years old), had mild motor deficits with an HFMSE at enrollment of 60/66 points. He was not under any treatment during the duration of the study. SMA02 (age range 51-55 years old) had the most severe motor deficit with HFMSE at enrollment of 38/66 points. During the study he was under Nusinersen. For safety reasons, he performed all the overground walking sessions with the use of the unweighing system (NxStem,Biodex), except the 6MWT. SMA03 (age range 26-30 years old) had moderate motor deficit with HFMSE at enrollment of 38/66 points. During the study he was under Nusinersen. A previous surgery on his right hip affected his gait. Thus all kinematic analysis was focused on his left leg.

### Safety

We meticulously documented all adverse events and promptly reported them to both the Data Safety and Monitoring Board (DSMB) and the Institutional Review Board (IRB) for a thorough evaluation of their potential association with the delivery of electrical stimulation to the spinal cord. Fortunately, all participants successfully completed the protocol without encountering any serious adverse events. The only non-serious adverse events were falls during the execution of exercises or during breaks between exercises that did not result in injuries (each participant fell once). None of these falls sustained any injuries. Risk mitigation strategies included the use of straps, harnesses, and the continuous presence of a physical therapist (PT) during exercises. This precautionary approach is crucial to ensure the safety of participants and underscores the importance of implementing robust measures to minimize the risk of falls during the course of the study.

### Biophysical model

The biophysical model was developed using NEURON ^68^.

### Motoneuron model

To model the dynamics of the membrane potential as a function of the different ion channels, we used the established Hodgkin-Huxley model. In this model, the membrane potential (*V)* is described by:

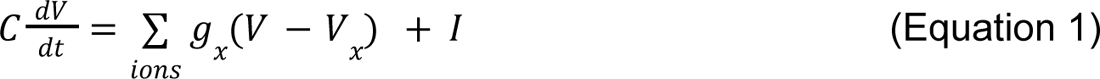

where *C* is the membrane capacity, *I* is current artificially injected into the neuron and *g*_x_ is the conductance of ion-channel *x* that, in general, depends on the membrane potential and ion concentrations. Following previous literature^35,38^, we included the following ion channels: delayed rectifier sodium (Na) and potassium (K-dr), N-like calcium (Ca-N), L-like calcium (Ca-L) and calcium-dependent potassium (K(Ca)). We used the motoneuron morphology and ion conductance described in a previous study^39^.

### SMA-affected motoneuron model

In a SMA mouse model, motoneurons exhibit impaired electrical properties. Specifically, SMA-affected motoneurons display reduced repetitive firing rate along with hyperexcitability, characterized by higher input resistance^5,6^. The impaired repetitive firing rate is a consequence of the reduction in the expression of the delayed rectifier potassium channel Kv2.1. Following these results, we blocked the delayed rectifier potassium channel (K-dr) by decreasing its conductance to 5% of its original value. With this change, we mimicked the impaired repetitive firing rate by lengthening the repolarization phase after an action potential (**Fig. 1d, e**). Instead, to reproduce the high input resistance we decreased the conductance of the leak and the sodium channel to 30% of their original values (**Fig. 1b**). These three modifications of ion-channel conductance mimicked the electrical properties of SMA-affected motoneurons.

### Network model

The biophysical model is composed of three populations of neurons: 1) Supraspinal neurons (N=200), 2) Ia sensory afferents (N=60) and 3) a pool of motoneurons (N=100). Both the supraspinal and the sensory afferents have excitatory connections to the MN pool (**Fig. 1f, Extended data Fig. 1**).

### Supraspinal neurons

We modeled them as poisson neurons to mimic natural firing rates from supraspinal sources.

### Ia sensory afferents

In contrast to the natural firing rate of the supraspinal neurons, Ia sensory afferents (N=60) fire synchronously and are controlled by SCS parameters. SCS had two parameters; the amplitude which controlled the number of fibers recruited and the frequency which controlled their firing rate.

### Motoneuron pool

In contrast to the supraspinal neurons and the Ia sensory afferent, we modeled the membrane dynamics of the motoneurons using Equation 1. Mimicking results from experiments in a SMA mouse model^5,6^, we modeled a SMA-affected motoneuron pool with two parameters; the percentage of motoneuron death and the percentage of surviving motor neurons to be SMA-affected (**Extended data Fig. 1a**).

### Excitatory Synapses

Each spike produced an instantaneous increase in current (*I*, Equation 1) followed by an exponential decay with τ = 2 *ms*. All supraspinal neurons and Ia-afferent fibers were connected to all motoneurons and their synaptic weights were randomly sampled from a gamma distribution. The parameters of the gamma distribution were adjusted to replicate the mean and variance of excitatory postsynaptic potentials in experimental data (mean EPSPs = 212 µ*V*)^39,69^.

### EMG model

To simulate the EMG produced by the motoneuron pool, action potentials were convolved with damped sinusoidal waves with a normally distributed amplitude (1 ± 0. 2 *a. u*.), normally distributed exponential decay ( 7. 5 ± 1. 5 *ms*) and a period of 15 *ms*.

### Simulation of the motor evoked potentials (MEP) from transcranial magnetic stimulation (TMS)

We simulated the MEPs from TMS stimulation by sending synchronized action potentials from the supraspinal neurons. To generate the recruitment, we progressively activated more supraspinal neurons (from 0 to 24) until all motoneurons were recruited. We defined the recruitment of 24 supraspinal neurons as a simulated TMS amplitude of 100% because the entire motoneuron pool was recruited.

### Simulated sensory-driven monosynaptic reflexes

Similarly to the TMS experiment, we simulated the sensory-driven monosynaptic reflexes by sending synchronized action potentials from Ia-afferents. Because synaptic weights from Ia-afferent were reduced by the effect of SMA the number of necessary Ia-afferent fibers recruited to activate the entire motoneuron pool was higher (52 Ia-afferent fibers) than from the supraspinal neurons.

### Force model

To simulate the force from the spike trains of all the motoneurons in a pool, we adapted a previously proposed model^70^. Here, we summarize this model. The twitch force generated for a spike from neuron *i* at time *t* is modeled as:

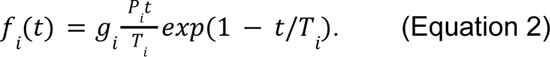

The peak twitch is defined as *P_i_ = exp(b • i)* and the contraction time is *T_i_ = 90(1/P_i_)^1/4.2^*. Finally, the gain, *g*_i_ was modeled depending on the normalized mean interspike interval of neuron *i* (*ISI*) and the contraction time *T*_i_: For *T_i_/ISI_i_ > 0.4*:

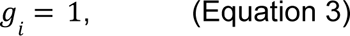

otherwise:

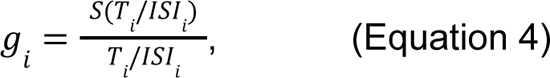

where *S*(*x*) is the sigmoidal function *S*(*x*) = 1 − *exp*(2*x*^3^).

Finally, the total normalized force is:

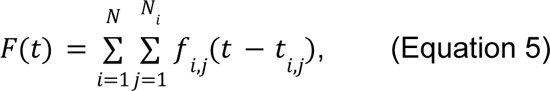

where *N* is the number of motoneurons and *N* is the number of spikes in neuron *i*. We always reported the force normalized with the maximum force produced by an intact motoneuron pool.

### Single-joint isometric torque

#### Torque measurement

The torques produced during maximum voluntary contraction were measured for hip flexion/extension and knee flexion/extension using a robotic torque dynamometer (HUMAC NORM, CSMi). To measure torque, the robot’s manipulandum was positioned and held at a fixed angle. Participants were asked to apply their maximum force while flexing or extending the specified joint for a sustained period of 5 s (SMA01) and 2 s (SMA02 and SMA03) followed by a 30s break. This procedure was repeated three times to complete a set. In general, participants completed two sets for each condition with 90 seconds of rest between sets. For each joint, the system was configured such that the joint was at a nominal and comfortable angle and that it was aligned with the manipulandum’s center of rotation. To isolate single-joint function, participants were constrained with straps across the shoulders and additional straps bracing relevant areas for each joint configuration of the HUMAC NORM. We used the HUMAC NORM’s suggested configurations. To measure knee extension/flexion participants were seated with their knee at a fixed position of 90° of flexion. For hip extension participants were in a supine position with their hips at 90°. For hip flexion, SMA01 was in a prone position with their leg extended back at 20°, a position where participants do not need to push their leg against gravity. For safety reasons, we measured SMA02 and SMA03 hip flexion in a supine position with their hip at 90°, the same position as hip extension.

We used the analog output of the HUMAC NORM to record the torque during each trial (smoothed it with a moving average of 100 ms). For each trial, we subtracted the weight of the limb recorded 4 seconds before the maximum voluntary contraction started. Measurements below 4 Nm were not reliable due to the biomechanical noise of our system. This was particularly important for SMA02 where we could only measure torque in his right hip. Instead, for SMA01, we could measure all joints and for SMA03 all joints except knee extension. (**Fig. 3, Extended Data** Fig. 4).

We tested the maximum torque produced by participants at the knee and hip extension/flexion pre-implant, post-explant and during the 4 weeks of SCS (knee extension/flexion two times per week, hip extension/flexion at least twice). To start every session participants completed 2 sets of 3 repetitions without SCS to track long-term therapeutic changes in torque across weeks. Then we turned SCS ON and repeated the same sets and repetitions. During the first and second weeks, we manually optimized the SCS parameters to maximize torque production for the hip and knee joints. During the sessions, we also tested sham configurations and other SCS parameters.

#### Analysis of fatigue in single-joint isometric torque

We used a multivariable linear regression analysis to control for fatigue across repetitions of maximum voluntary contraction repetitions. In this analysis, we used the following regressors: day (day), the number of maximum voluntary contractions already performed in that day (rep), the presence of SCS (SCS), the repetition number inside each set of repetitions (RepSet), the interaction between the repetition number and SCS (RepxSCS) and a constant which captures the torque before the study. We defined the variable day as the number of days from the start of the torque improvement. To find this day, we added an offset in days, setting all days before it to 0, and systematically optimized the parameters by adjusting this offset. We defined the day when the torque began increasing as the offset used in the optimization with better R^2^ value. Parameters were optimized using ordinary least squares. We performed this analysis on single-joint torques with enough data (e.g. tested it at least 5 of the days during the 4 week implant period). This analysis revealed that for SMA01 the repetition number was significant indicating that the maximum torque produced decreased with the repetition number. For this reason in SMA02 and SMA03, we reduced the time of maximum voluntary contraction from 5 to 2 s. Indeed, for SMA03 the repetition regressor was not significant. Interestingly, we found that SCS was able to increase the maximum torque of SMA01 during fatigued trials in right knee extension (SCS interaction between repetition and SCS significant) but not at the beginning of the session (SCS regressor not significant, **Extended data Fig. 4b**). For this reason, in **Fig. 3d** for SMA01 right and left knee extension, we compared the torque with and without SCS when SMA01 was already fatigued, the last 12 trials in sessions with more than 20 maximum voluntary contraction repetitions.

#### Analysis of the assistive effect in maximum torque

In every session, we measured the torque produced without SCS and repeated the same measurements with SCS. We computed the percentage of increase with SCS and the confidence intervals with bootstrap (N=10,000) (**Fig. 3d**).

#### Analysis of the therapeutic effect in maximum torque

To reduce the variability of maximum torque production across days, we defined the data recorded pre-implant and week 1 as pre-study, while week 4 and post-explant as post-study. We computed the percentage increase in torque without SCS pre vs post-study and used bootstrap to compute the confidence interval and level of significance. In this case, we used a one-sided test because our hypothesis is that SCS, in the long-term, will improve maximum torque production (**Fig. 3e-g, Extended data Fig. 4a**)

### Kinematic Curves, Gait Variables and EMG Envelope

Once per week during the study, participants were instructed to walk back and forth at a comfortable speed in the gait laboratory. We combined trials with and without SCS and we took breaks between laps to prevent fatigue. We placed 16 retroreflective markers on the surface of the skin and used the Vicon Motion Capture system with 12 infrared cameras (100 Hz frame rate) and 3 optical cameras (60 Hz frame rate) to track the movement of the participants. We recorded the EMG activity of 7 different muscles in each leg (Supplementary Information) Joint angle motions were computed using the Nexus biomechanical software (VICON). We developed custom software in MATLAB (MathWorks) to detect gait events (foot strike/foot off). We segmented the data between two consecutives foot strikes of the same leg and defined this as one gait cycle. We high pass filter (first-order Butterworth high pass filter, cutoff frequency of 0.01Hz) and segmented the data to remove offsets between sessions due to slight changes in the placement of the markers. Finally, we resampled each gait cycle to have 101 samples.

We computed the range of motion (ROM) of each joint angle across the gait cycle and analyzed the percentage change for the assistive (SCS ON vs OFF, same day) and therapeutic effects (wk1 vs wk4, SCS OFF). Gait variables (step length, step height and stride velocity) were computed based on the 3D trajectory of the heel markers. Specifically, step length was determined as the distance covered in the direction of motion between two consecutives foot strikes of different legs; step height was measured as the maximum height during each step, and stride velocity was calculated as the ratio of stride length (distance between consecutive foot strike of the same leg) to stride duration. For SMA01 and SMA02 we computed these parameters for both legs. However, for SMA03, we only used the data from the left leg because the gait of their right leg was affected by a surgery previous to this study. Gait variables for each subject were normalized for their maximum median value across conditions (i.e. different weeks or SCS ON vs OFF).

EMG data were synchronized with the kinematic data, high pass filtered (sixth-order Butterworth high pass filter, cut-off frequency of 50 Hz), full-wave rectified and smoothed (first-order Butterworth low pass filter, cut-off frequency of 4 Hz). Each envelope was then segmented across gait cycles, resampled to 1001 samples and normalized according to the maximum signal amplitude. To evaluate the change in muscle activation, we evaluated the percentage change in root mean square (RMS).

### Kinematic during treadmill walking

Throughout the experimental sessions, participants were directed to walk on an AlterG® antigravity treadmill. Under continuous supervision by a Physical Therapist, SMA01 was guided to walk, emphasizing hip flexion, at a comfortable speed with and without SCS. We captured kinematics using a GoPro® Camera and analyzed the data with Deeplabcut^71^, a neural network designed for 2D and 3D markerless pose estimation based on transfer learning. A subset of video frames per condition was selected by Deeplabcut to ensure diversity of movement. Subsequently, we manually labeled the ankle, metatarsal, and toe in each of these frames. Using these labeled frames, we trained a network to analyze all the videos and extract kinematic traces for each labeled body point. Specifically, utilizing the trajectories of the metatarsal marker, we calculated the step height for each gait cycle (**Extended Data** Fig. 5e-g).

### Single motoneuron firing rate analysis

#### Motoneuron decomposition

We recorded high density surface electromyography (HDEMG) from knee extensors (i.e., Rectus Femoris (RF)) and flexors (i.e., Biceps Femoris (BF)) while the participants performed two sets of three Maximum Voluntary Contraction (MVC). An 8×8 channel flexible HDsEMG grid electrode with 8.75 mm distance between electrodes was placed over the RF and BF muscles, respectively. Conductive gel was used to reduce the skin-electrode impedance. At the end of each experimental session the edges of the grid were traced on the participants’ skin using a skin marker to ensure consistent grid placement throughout the different phases of the clinical study (i.e., pre-implant (PI); i-th week after the implant (Wi); post-explant (PE)). EMG recordings were acquired in monopolar configuration, with the reference electrode placed over the patella, using the TMSi Saga 64+ high density amplifier at a sampling rate of 4 kHz. We then decompose the HDsEMG recordings into the spike train of individual motoneurons using DEMUSE tool software which exploits the convolution kernel compensation (CKC) method^72^ (**Extended Data** Fig. 7a). The results of this automatic decomposition were manually edited following the standard procedure already described^73,74^. Only motoneurons with pulse-to-noise ratio ≥ 30 dB^75^ were selected for further analysis.

#### Rescued motoneurons identification

To assess changes in the motoneuron firing properties during the clinical study, we compared the peak firing rate of motoneurons without SCS between the first and last sessions where we recorded HDsEMG (week 2 vs week 4 in SMA01, week 1 vs week 4 for SMA02 and pre-implant vs post-explant in SMA03). We created a unique set of motoneurons per experimental session, motoneurons were tracked across the two sets of MVC using the motor unit filter transfer method^76^. We calculated the motoneuron firing rate in a 100 ms overlapping window and subsequently smoothed with a 100 ms moving average window to obtain the smoothed discharge rate (SDR). A post-study motoneuron was considered functionally rescued when its mean peak firing was above the 99.73% CI of the mean peak firing rate among the pre-study motoneurons (**Extended Data** Fig. 7b). We computed this CI using bootstrap (N=10,000). To test the hypothesis that functionally rescued motoneurons have a higher firing rate, we performed a one-sided significance test using bootstrap (N=10,000).

#### Comparison SCS ON vs SCS OFF

To evaluate changes in motoneuron firing rate mediated by SCS, we tracked the same motoneurons in trials with and without SCS using the motoneuron filter transfer method^76^. We used the motoneurons that we could track in both conditions to compare the mean peak firing rate with and without SCS. To test the hypothesis that motoneurons have higher firing rate in SCS ON vs OFF conditions, we performed a one-sided significance test using bootstrap (N=10,000).

### Transcranial Magnetic Stimulation

Magnetic stimulation was applied to M1 by a magstim 200 through a figure-of-eight coil (70 mm loop diameter, D70) with single, monophasic pulses (The Magstim Company Ltd, Whitland, UK). Measurements were taken using the 10-20 system to identify scalp locations corresponding to the vertex (Cz) and and ∼1 cm lateral of the vertex line in line with the tragus of the ear contralateral to the target muscle. The optimal scalp site was determined by stimulating this location and moving the coil in increments anterior-posterior/medial-lateral directions following a 5×5 cm grid with 1 cm spacings. The coil was also rotated to identify the optimal angle relative to the mid-sagittal plane. The location and angle that produced stable MEPs at the near threshold stimulator outputs was set as the optimal location and recorded by frameless, stereotaxis neuronavigation system (Brainsight, Rouge Research Inc,m Montreal, Quebec, Canada). Coil location and orientation were adjusted by the experimenters throughout the duration of each experiment using feedback provided by the neuronavigation system. At the optimal targets stimulation was applied for 5 or 10 pulses with an inter-stimulus interval of at least 5 seconds. Stimulation intensity was stepped from 60% of stimulator output to 100% at increments of 5%. This test was performed pre-implant, third week of trial, post-explant and 6 weeks follow-up. SMA01 could not perform the follow-up TMS session.

### Recruitment Curves

To evaluate the specificity of SCS in recruiting individual motor pools, recruitment curves were Performed on representative contacts of the lead and optimal configurations found throughout testing. We used a wireless EMG system (Trigno, Delsys Inc.) to record compound muscle action potentials (CMAPs) elicited by SCS pulses. SCS was delivered at 1-2 Hz on one electrode at a time with gradually increasing current amplitude while simultaneously recording CMAPs from all muscles. The peak-to-peak amplitude of the SCS-induced CMAPs were measured, one for each stimulus amplitude, and normalized to the maximum amplitude recorded on that muscle across all measured trials.

### Spinal reflexes during passive joint movements

The Humac Norm Cybes (HUMAC NORM, CSMi) was used to impose passive joint movements with a sinusoidal profile of fixed amplitude and frequency, while continuous SCS was delivered to produce motor responses in the muscles spanning this joint. Subjects were asked to relax, neither to resist, follow, nor facilitate the movements. Muscle responses were recorded using the wireless EMG electrodes. SCS parameters and the amplitude of the movement were set depending on subject-specific constraints (**Fig. 2e, Extended Data** Fig. 3). Joint kinematics were recorded with a sampling frequency of 8000 Hz. SCS parameters were chosen in order to recruit the targeted muscles. SCS was delivered in frequencies of 1-2 Hz. SCS amplitude was set in order to induce consistent muscle responses across the targeted muscles. For each participant 15 minutes of recording was performed. To quantify the modulation of muscle responses during the passive movements, we extracted the timing of each SCS pulse with the recorded stimulation pulses. We then extracted the muscles responses and grouped them according to the phase of the cyclic movement (n = 10 bins) (**Extended Data** Fig. 3). We used bootstrap to compute the 95% CI of the peak to peak responses (N=10000).

### X-ray Imaging

X-ray images were acquired at weekly time points in both the axial and sagittal views to ensure the stability of lead position

### Functional Imaging Data

#### Active Task

During each scan session, participants completed three 6 minute runs composed of 16 second blocks of active knee extension where the knee is extended and relaxed at a rate of 0.5 Hz. One run consisted of 9 active extension blocks.The aim was to target the quadriceps and hamstring muscles. A custom software implemented in PsychToolbox v3.0.17 allowed instructions and repetitions to synchronize with the MRI acquisitions. Instructions were displayed on a screen (fixation cross ‘+’ during rest blocks and text indicating activity blocks), while auditory cues were used to signal changing blocks. Only the participant’s right leg was tested.

#### fMRI data Processing

The fMRI pre-processing utilized the FMRIB Software Library (FSL) v6.06^77^ and the Spinal Cord Toolbox (SCT) v6.1^78^. The first five volumes of each run were removed to allow for T1 equilibration effects^79^. The volumes of each run were averaged to create a mean functional image and was used to automatically detect the centerline of the spinal cord^80^. A cylindrical mask (diameter of 30 mm) was generated along the centerline to prevent the inclusion of regions moving independently from the spinal cord. Initial steps involved a two phase motion correction procedure. Using three-dimensional rigid body realignment (spline interpolation and least square cost function) all volumes within a run were registered to the mean image using FSL’s MCFLIRT^79^. Acknowledging the non-rigid nature of the spinal cord, two-dimensional slice-wise realignment (spline interpolation and least square cost function) was performed with the mean functional image as the target^78^. Finally, all runs corresponding to the same session were aligned to the first run of the session using three-dimensional rigid body realignment (spline interpolation and least square cost function). Motion scrubbing was employed to identify outlier volumes using FSL’s outlier detection tool (DVARS metrics: root mean square intensity difference of volume N to volume N+1) within the spinal cord using a box-plot cut-off (75th percentile + 1.5 x the interquartile range)^81^. Both the cerebrospinal fluid and the spinal cord were automatically segmented (with manual corrections when necessary) using the Spinal Cord Toolbox (SCT)^82^ from the mean motion corrected functional images and the T2 anatomical images. Noise regressors were generated using FSL’s physiological noise modeling tool on the acquired cardiac and respiratory signals, with a procedure based on the RETROICOR^83^. Low and high order Fourier expansions were used to model the physiological signals^84,85^. Accordingly, we generated 32 noise regressors using the physiological noise modeling (PNM) tool from FSL, along with an additional regressor corresponding to the cerebrospinal fluid signal (10 % most variable cerebrospinal fluid voxels).

Using the Spinal Cord Toolbox, functional images were coregistered to the PAM50 image with non-rigid transformations using the conus medullaris as landmarks to create functional-to-template warping fields. The 33 physiological noise regressors, motion correction parameters (x and y), and motion outliers were regressed from the fMRI time-series using FSL’s Expert Analysis Tool (FEAT). The resulting residuals were spatially smoothed, volume by volume, using a 3D Gaussian kernel (with full width half maximum (FWHM) of 2 x 2 x 6 mm^3^, along the centerline of the spinal cord, to preserve consistency at the anatomical level. Spinal segments L1 to S2 were identified using the high-resolution structural MRI. The L1 dorsal root was identified from its entry region in the spinal canal (entering just below the L1 vertebra) until the region where it innervates the spinal cord, which defines the L1 spinal segment. The more caudal segments (L2 - S2) were identified by following the dorsal roots along the rostrocaudal axis.

#### fMRI data analysis

The resulting smoothed residuals were then entered into a first-level statistical analysis with local auto-correlation correction^86^. As explanatory variables, the timings of the task (block design) were convolved with the three optimal basis functions using FMRIB’s Linear Optimal Basis Set^87^, with the second and third waveforms orthogonal to the first waveform. The resulting parameter estimates for the three runs were passed through a fixed-effects model to obtain the second level analysis (subject level and task specific) activation maps. Activation maps were transformed to the PAM50 template using the functional-to-template warping fields to allow for comparison across sessions. Z statistic images were presented as uncorrected (Z > 2.3, p < 0.01). These results were then registered to the respective anatomical image to assess their spatial distribution with respect to spinal segments.

### Clinical Evaluations

#### Revised Hammersmith Functional Scale (RHFS)

The Revised Hammersmith Scale (RHS) is a clinical outcome measure specifically designed to assess motor function in patients with Spinal Muscular Atrophy. The Hammersmith Functional Motor Scale Expanded (HFMSE) was developed for patients with SMA who are ambulatory. The HFMSE consists of 33 items that are scored either 0, 1 or 2. A score of 2 is assigned to participants who achieve the motor task without any compensatory strategies. Attempted movements or items achieved with compensation are scored a 1. A score of zero is assigned to those unable to perform the task. The same licensed therapist administered and scored HFMSE tests pre and post-study.

#### Six-minute walk test

The Six-Minute Walk Test (6MWT) is a sub-maximal exercise test of endurance and aerobic capacity. Participants are instructed to “walk as far as possible for 6 minutes”. The distance walked over the course of six minutes is measured in a structured environment. The 6MWT is routinely implemented to assess changes in basic mobility in patients with SMA. We compute the fatigability index as the percentage change in velocity between the first and last lap. The same licensed therapist administered and scored the 6MWT pre and post-study.

### Statistics

#### Bootstrapping

All confidence intervals in this manuscript were performed using bootstrap with N=10,000. To compute p-values, we computed the probability to obtain the observed result or a more extreme value under the null-hypothesis distribution created with bootstrap.

#### Boxplot

The central mark denotes the median, while the bottom and top edges of the box indicate the 25th and 75th percentiles, respectively. Whiskers extend to the most extreme data points not considering the outliers.

## Extend Data Tables

**Extended Data Table 1:** Clinical assessments. HFMSE and 6MWT measured pre-implant, after 4 weeks of SCS therapy and at 6 weeks follow up. In parenthesis changes over previous measures. All tests executed with SCS OFF. 6MWT dropped back to pre-implant at 6 weeks follow up in 2/3 participants.

|  | HFMSE pre | HFMSE post | HFMSE follow up | 6MWT pre | 6MWT post | 6MWT Follow Up |
| --- | --- | --- | --- | --- | --- | --- |
| SMA01 | 60 | 61 (+1) | 61 | 520 | 552 (+32 m) | 500 m (-52 m) |
| SMA02 | 38 | 40 (+2) | 43 (+3) | 72 | 92 (+20 m) | 93.5 m (+1 m) |
| SMA03 | 49 | 49 | 49 | 102 | 126 (+24 m) | 108 m (-18 m) |

## Extended Data Figures

**Extended data figure 1:**
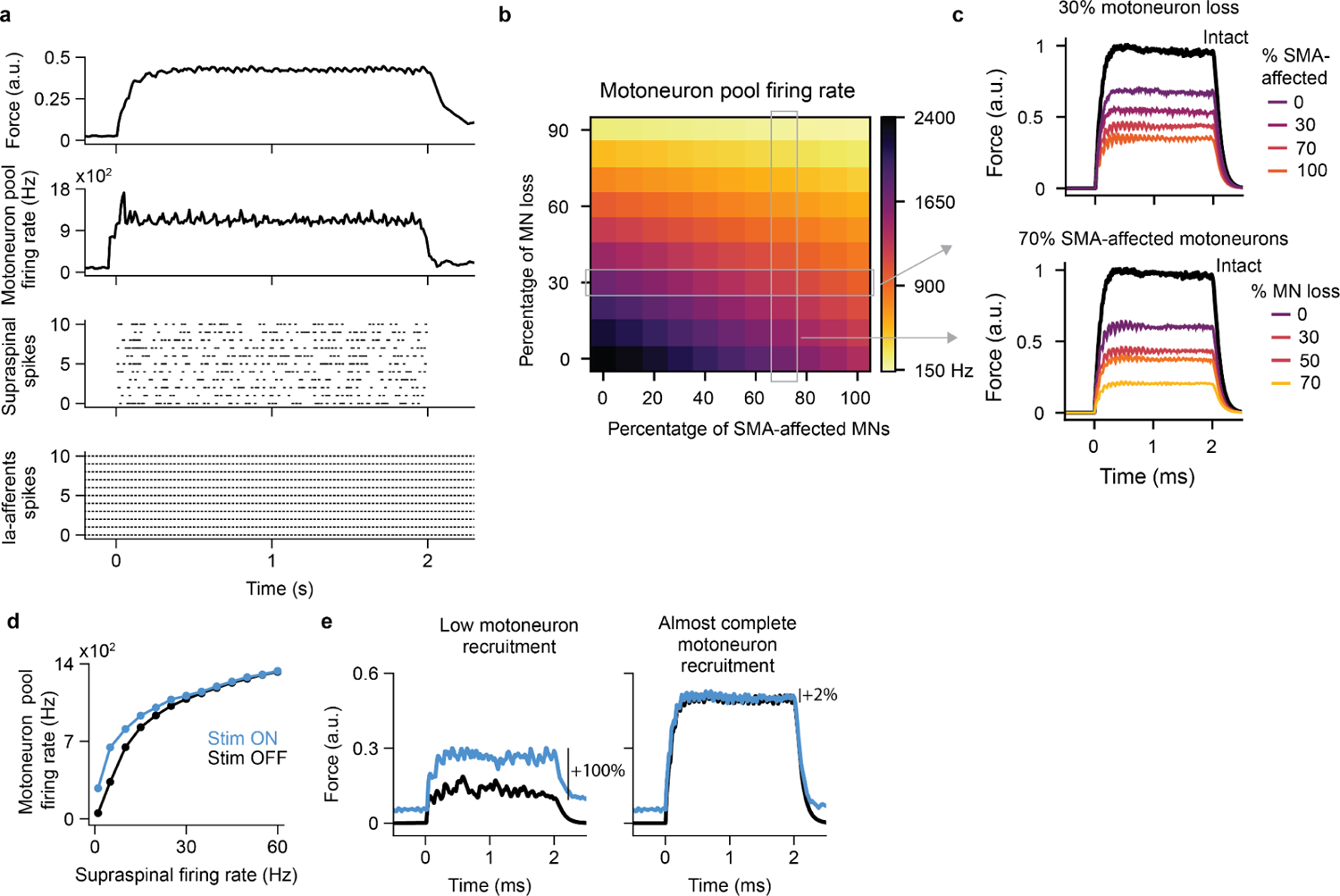
Biophysical model. **a,** Example of 2 second isometric force produced by a SMA-affected motoneuron pool simulated with the biophysical model. From bottom to top: Raster plot of 11 Ia-afferent fibers recruited by SCS at 40 Hz. Note that spikes are completely synchronized because they are triggered by SCS pulses. Supraspinal neurons with natural firing rate (Poisson neurons); for visualization propose, we only show 11 neurons but the supraspinal population has 200 neurons. Motoneuron pool firing rate with 30% of motoneuron loss and 50% SMA-affected motoneurons. Simulated force produce by the motoneuron pool normalized by the maximum force produced by an intact motoneuron pool. **b,** SMA-affected motoneuron pool firing rate as for different simulated percentage of motoneuron loss and SMA-affected motoneurons. **c,** Simulated force for different percentages of motoneuron loss and SMA-affected motoneurons. **d,** Motoneuron pool firing rate as a function of the supraspinal firing rate with SCS ON and OFF. The biophysical model predicted that the effect of SCS should be more prominent on low than high supraspinal inputs indicating that if all motoneurons are recruited at their maximum firing rate, there should be no effect of SCS. Thus, the effect of SCS at maximum voluntary contraction will depend on the participants’ ability to recruit the resources of their motoneuron pool. SCS could give access to motoneurons that are not recruited or recruited with a submaximal firing rate. **e,** Simulated force with a low supraspinal input (5Hz) and very high supraspinal input (50 Hz).

**Extended data figure 2:**
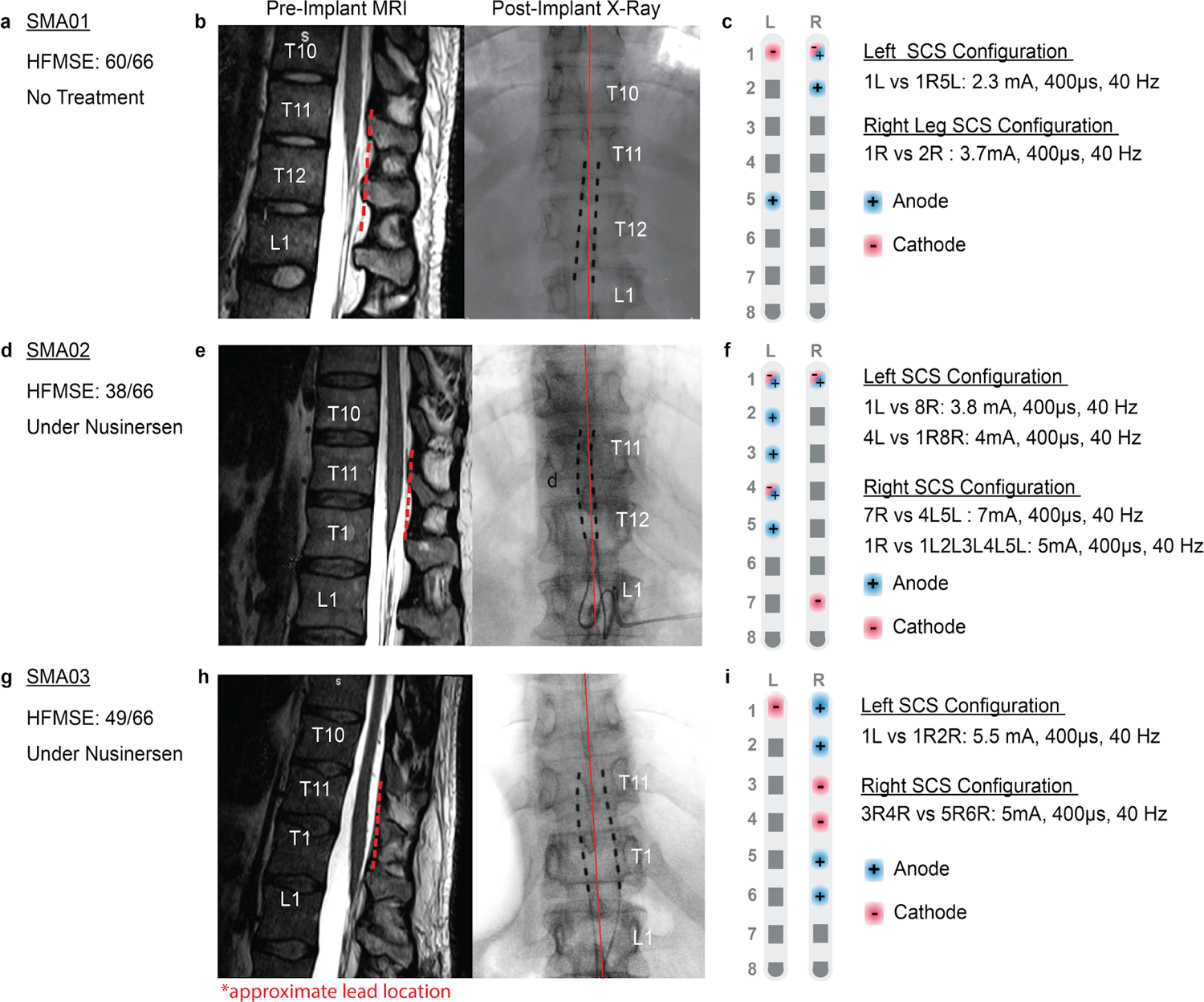
Implants and SCS configuration. **a, d, g**. For each study participant demographic information, including ethnicity, age, score at the enrollment on the Hammersmith Functional Motor Scale Expanded (HFMSE), and detail on any treatment used. **b, e, h**. Left: pre implant T2 MRI, with the approximate lead location in red. Right: X-ray post implant, with the bilateral leads in black. **c, f, i**. SCS parameters used during walking. We used a program targeting the right and a program targeting the left leg muscles. Each program applied a fixed amplitude, frequency and pulse width for each SCS pulse.

**Extended data figure 3:**
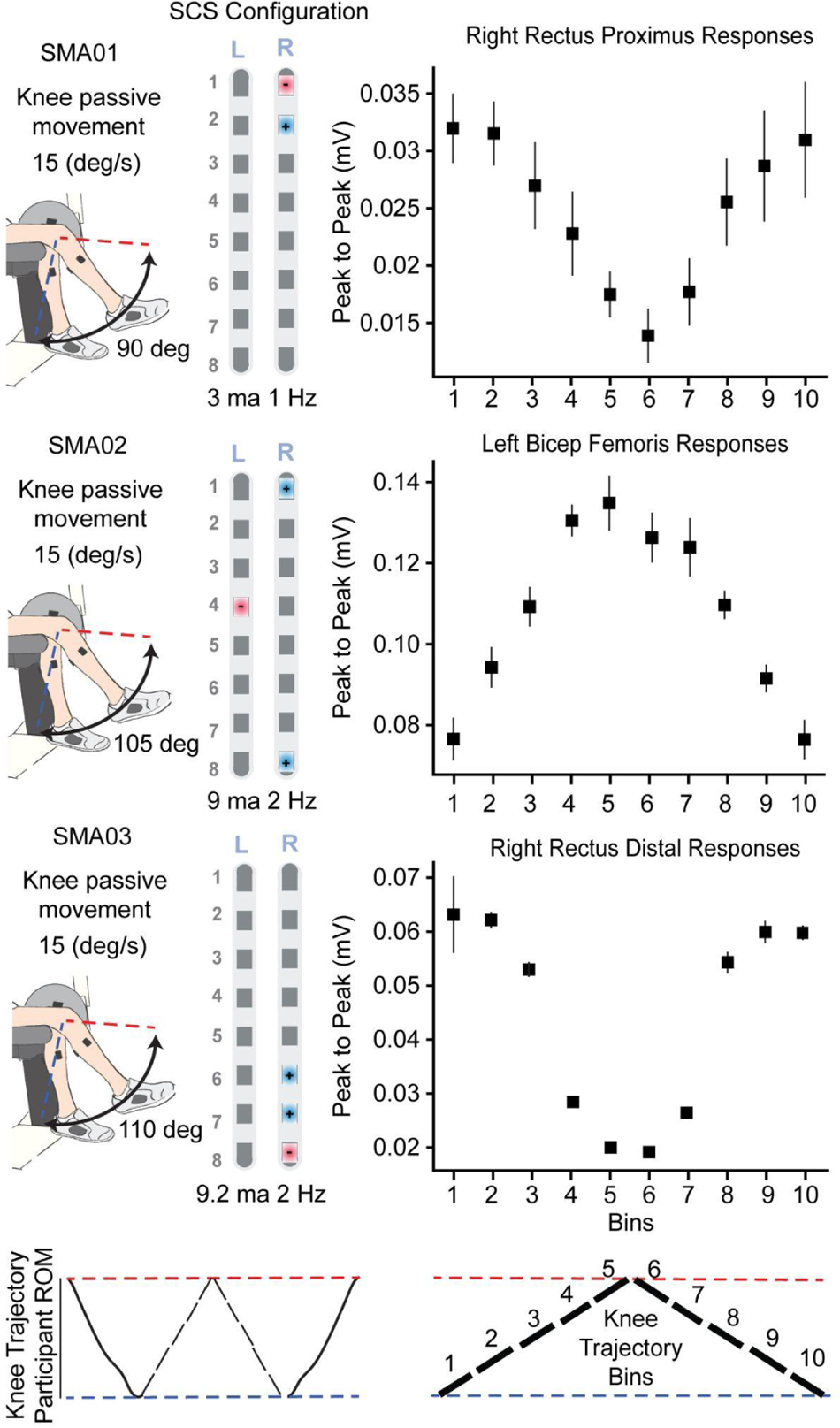
Effect of SCS on the natural modulation of sensory-motor circuit during passive movements. Left: Configurations of the experimental set ups for all participants and the SCS parameters used (anode blue, cathode red). Bottom: The cycle of joint oscillations was divided into 10 bins of equal duration during which the muscle responses were extracted and regrouped. Right: Muscle responses were used to compute the peak to peak of each bin. Plots are reporting the mean and 95% CI of the peak to peak responses. All error bars indicate the confidence interval, computed with bootstrap (N=10,000).

**Extended data figure 4:**
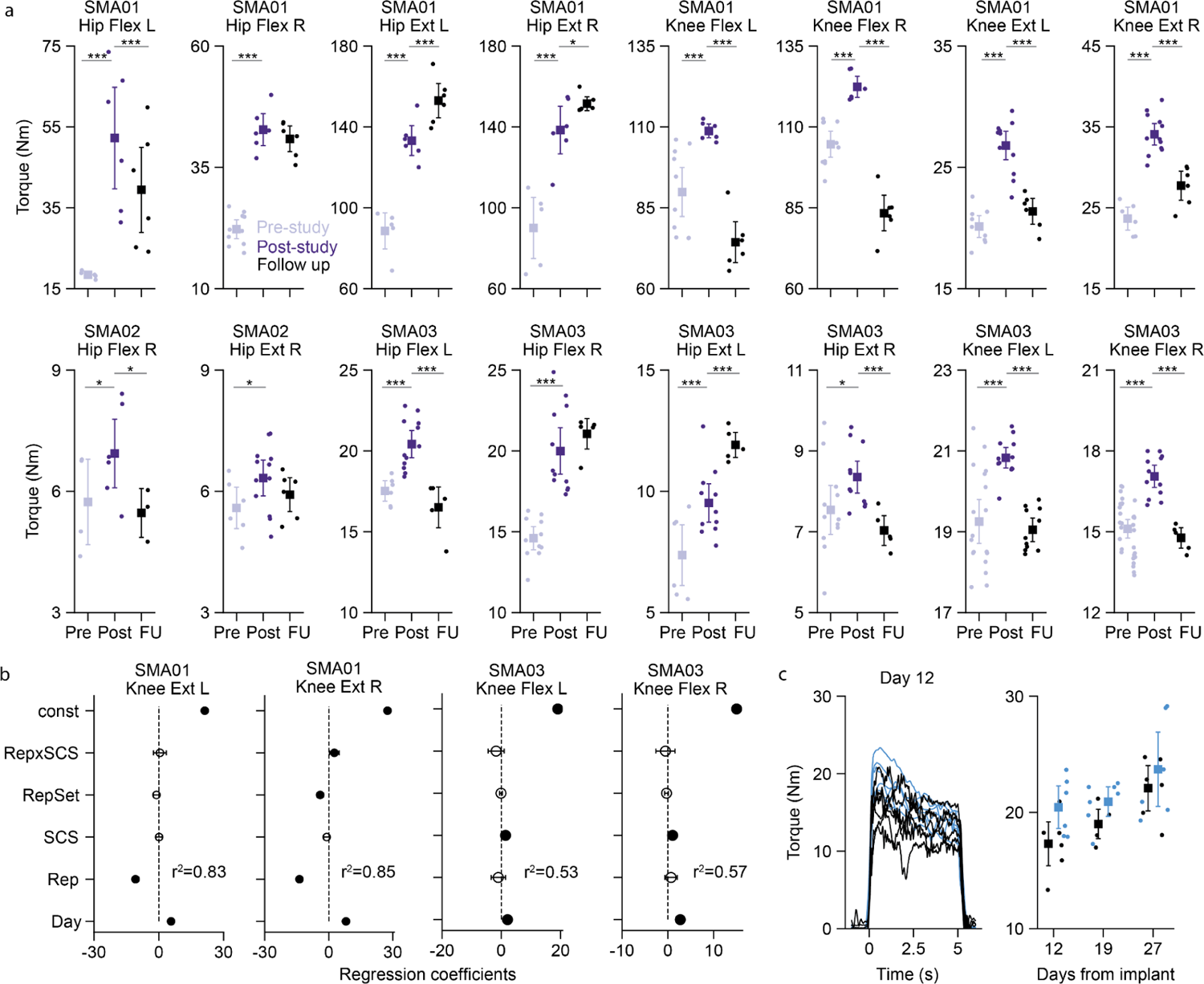
Therapeutic effects in torque and fatigue analysis. **a,** Maximum strength in hip and knee for all participants at pre-study, post-study and follow up ( 6 weeks after post explant). To reduce the day to day variability in our measures of maximum torque, we used data from pre-implant and week 1 to evaluate pre-study torque. Similarly, the post-study torque was computed using the data from week 4 and post-study. Error bars show the 95% confidence interval. One-sided significance test, p<0.05 (*), p<0.001(***). **b,** We used a multivariable linear regression analysis to study the effect of fatigue in our measurements of torque (see methods). For SMA01, we found that the largest regressor was the repetition indicating that SMA01 was fatiguing after only a few repetitions of maximum voluntary contractions. Given that we always started with SCS OFF trials, our results could be underestimating the effect of SCS (Fig. 3d). Indeed, we found for SMA01 right knee extension, the interaction between the SCS and the repetition number was significant indicating that SCS increased maximum torque when SMA01 was fatigued. In SMA03 we reduced the trial duration time from 5 to 2 seconds and we found that the repetition number had no impact indicating that direct comparison between stim off and on trial is probably not affected by the repetition number. **c,** Maximum torque at right knee extension produced by SMA01 in fatigued trials (after more than 12 trials of maximum voluntary contraction).

**Extended data figure 5:**
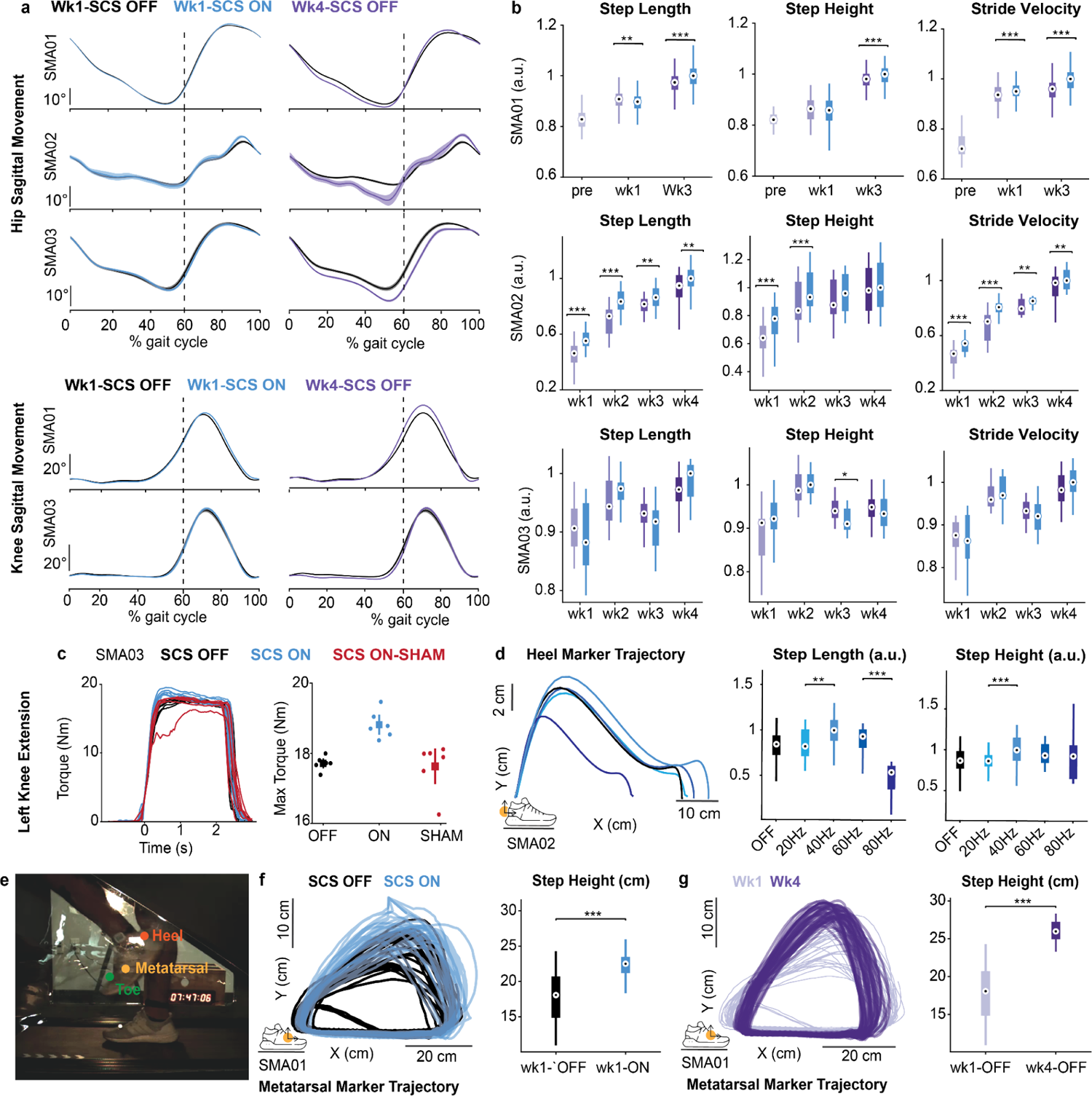
Gait and Sham. **a**. Hip and Knee kinematic curves spanning the gait cycle are compared between SCS-off and SCS-on conditions in week 1 and week 4 with SCS-off. Average curve and confidence interval bootstrap-derived (N=10,000). **b**.Gait variables, including step length, step height, and stride velocity, across weeks for both SCS-off and SCS-on conditions. Box plots, illustrating the median, 25th, and 75th percentiles, unveil the minimum and maximum data points, with outliers excluded. **c**. Left: Examples of isometric torque traces during maximum voluntary knee extension contraction are provided, showcasing performances with SCS, without SCS, and with sham SCS parameters. Right: Maximum torque during knee extension is compared across conditions: without SCS, with optimal SCS, and with sham SCS parameters. **d**. Left: mean heel marker trajectory across gait cycles while the participant experiences SCS with the same program but different frequencies. Right: step length and step height are computed under the same conditions. **e**. Setup for testing exaggerated hip flexion walking on the anti-gravity treadmill. **f**. Examples of metatarsal marker trajectory during exaggerated hip flexion walking are presented, with blue indicating stim-on and black indicating stim-off. On the left, step height is computed for each gait cycle. **g**. same to g, but considering stim-off conditions in week 1 and week 4. Statistical differences are consistently assessed using two-tail bootstrapping (N=10,000). Asterisks (*) indicate significant differences, with p-values: p < 0.05, p < 0.01, and p < 0.001, respectively.

**Extended data figure 6:**
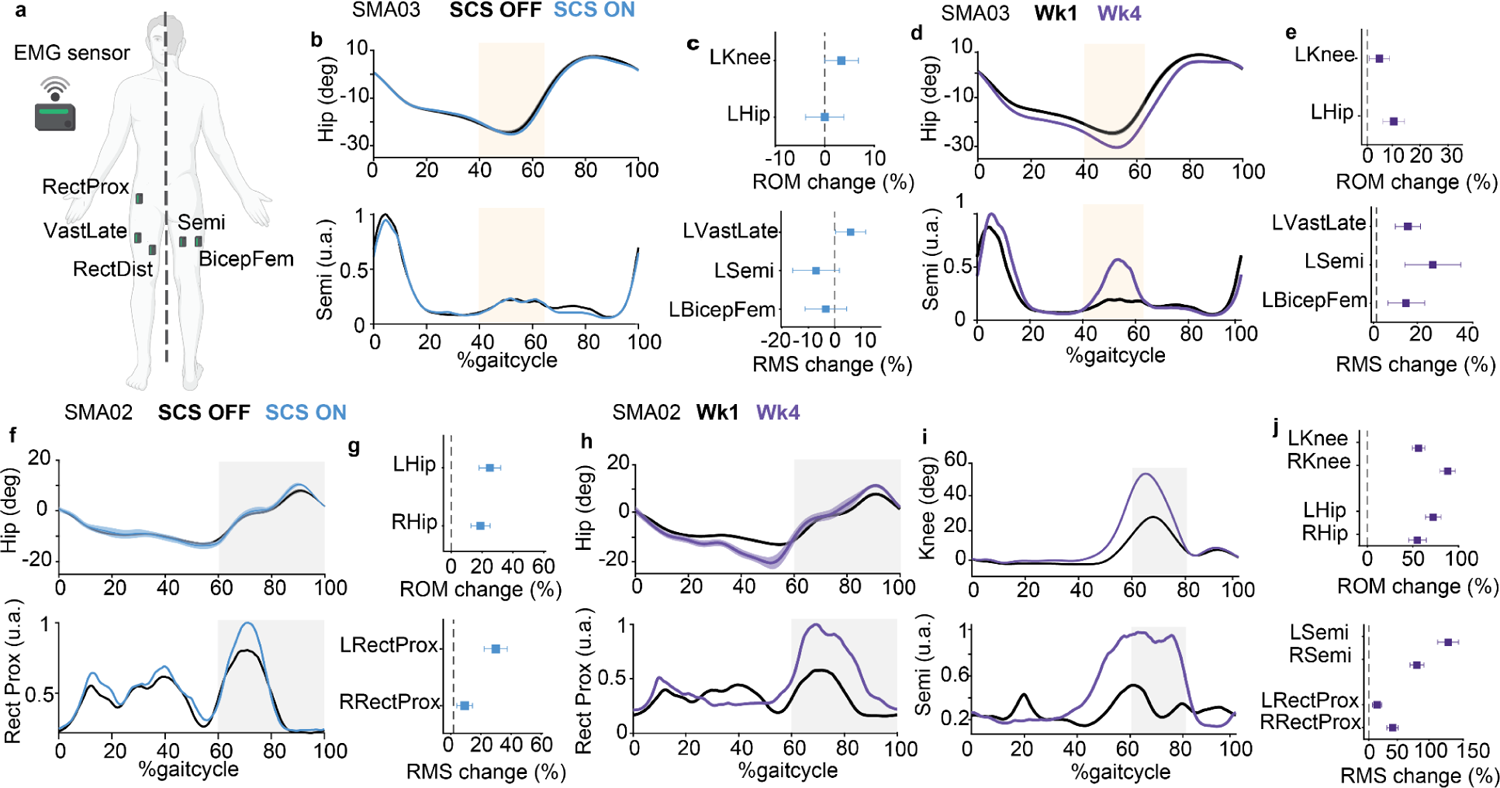
Increases in range of motion are paralleled by increases in EMG activity. **a**. Scheme depicting the placement of the EMG sensor during overground walking. **b-c** Assistive effects in range of motion (ROM) for SMA03. **b,** Top: Mean hip flexion/extension across the gait cycle with SCS OFF (black) and SCS ON (light blue). Bottom: Mean envelope of Semitendinosus (Semi) across the gait cycle with SCS OFF (black) and SCS ON (light blue). The gold shades highlight semitendinosus activation in the pre-swing, contributing to hip extension during the same phase of the cycle. **c.** Top: Change in ROM for the left hip and knee. Bottom: Change in EMG activity, computed as the root mean Square, for Semitendinosus (Semi), Vastus Lateralis (VastLate), and Biceps Femoris (BicepFem). Improvements in hip ROM should be correlated with the EMG activity at Semi and BicepFem (muscles involved in hip extension). Improvements in knee extension should be related to higher activation on the knee extensors. No change of EMG or ROM. **d-e** Therapeutic effect in ROM for SMA03. Improvements in hip and knee ROM are paralleled by an increase in EMG activity at hip and knee extensors **f-g**. Assistive effects in ROM for SMA02. The gray shades highlight Rectus Femoris proximal activation in the swing phase, contributing to hip flexion during the same phase of the cycle. An increase in hip ROM can be explained by an increase in EMG activity at the Rectus Femoris proximal (a proxy of EMG activity of the hip flexors such as the iliopsoas) **h-j.**Therapeutic effect in ROM for SMA02. Improvements in hip and knee ROM are correlated with increases in EMG activity of hip flexors (rectus femoris proximal) and knee flexors (Semi) No data is reported for SMA01 due to EMG acquisition corruption by noise.

**Extended data figure 7:**
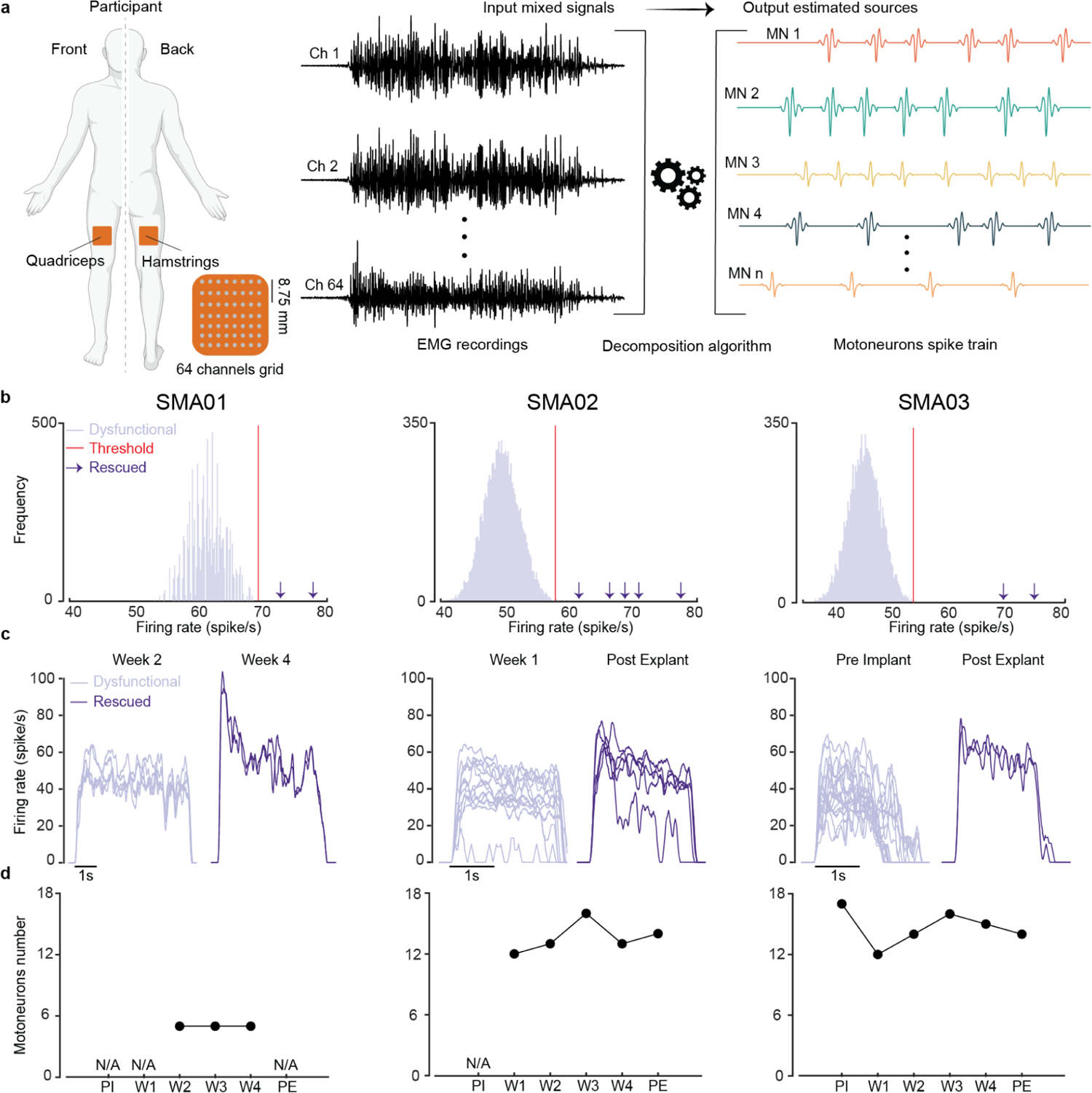
High Density EMG decomposition in single motor units spike trains. **a.** Pipeline to extract single motor units spike trains from high density surface electromyography (HDEMG). We used an 8×8 channel flexible grid to record muscle activity from both knee extensor (Rectus Femoris (RF)) and flexor (Biceps Femoris (BF)). Participants performed two sets of three isometric Maximum Voluntary Contractions (MVC) of knee extension and flexion. We used the convolution kernel compensation (CKC) method to decompose the EMG signals in single motor units spike trains. **b.** Rescued motoneuron identification: a motor unit was defined as rescued if its mean peak firing rate (across trials) was higher than the 99.73 % CI (red line) of mean peak firing rate across pre-study motor units. The 99.73 % CI was computed with bootstrap (N=10,000). **c.** Example of motor units smoothed discharge rate (SDR) obtained during the 3rd maximum voluntary contraction. Each line represents the SDR of every motoneuron. Light and dark colors delineate pre and post-study motoneurons, respectively. **d.** Circles illustrate the number of identified motoneurons over the different phases of the study. (i.e., PI = pre-implant; Wi = i-th week after the implant; PE = post-explant).

**Extended data figure 8:**
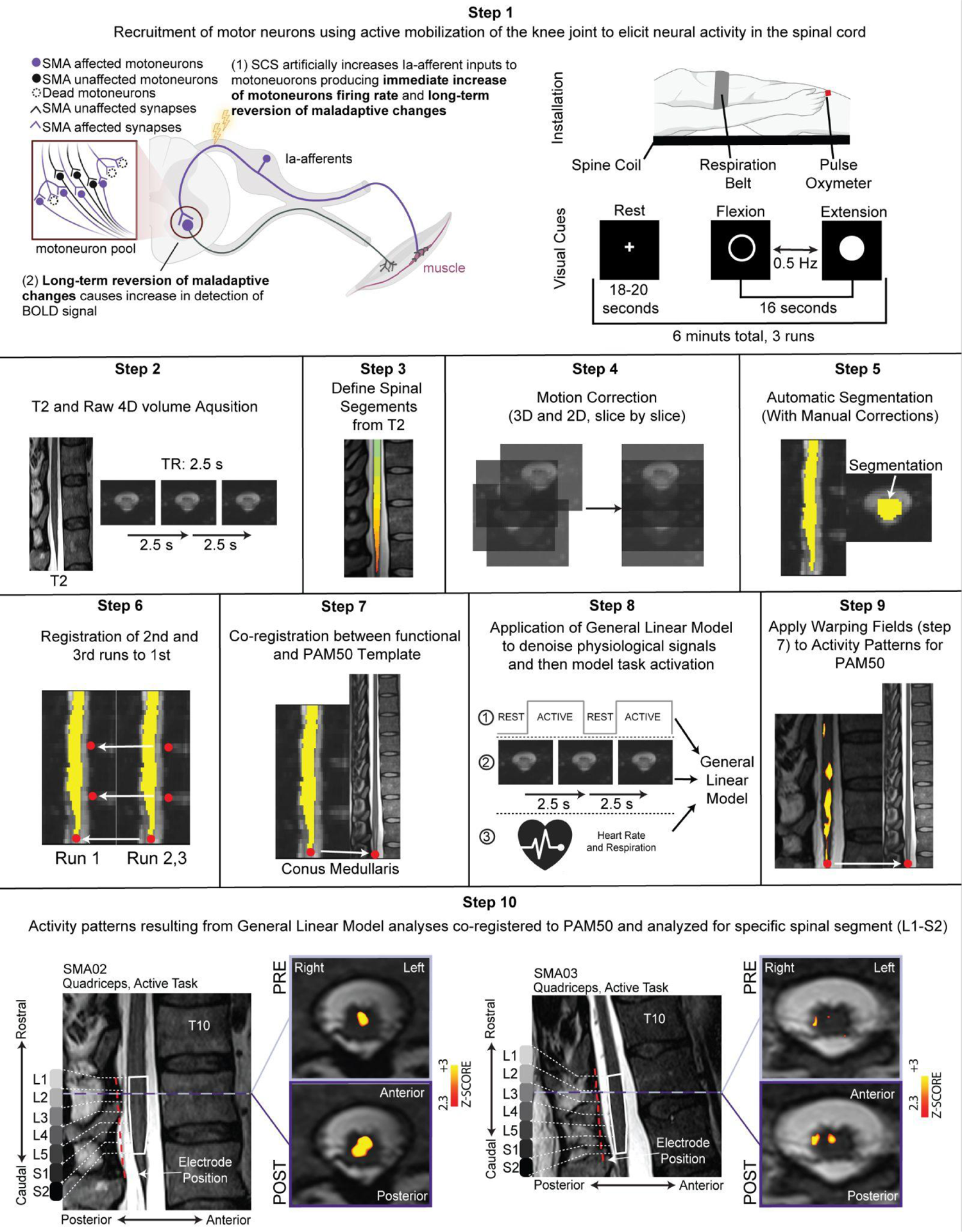
fMRI to detect spinal motoneuron function. **Step 1**, Acquisition of functional MRI from the spinal cord in response to the recruitment of motor neurons from specific leg muscles. The motor neurons are recruited by actively extending and flexing the knee joint. Three runs are acquired for each participant. Only the right leg muscles are tested. Instructions were displayed on a screen (fixation cross ‘+’ during rest blocks and text indicating activity blocks), while auditory cues were used to signal changing phases. In addition to the functional volume series, T2 anatomical images and physiological (heart rate, respiratory) signals are acquired. **Step 2**, Raw fMRI volume series are repeatedly acquired every 2.5 s (TR) in functional runs lasting about 6 minutes. **Step 3**, Spinal segments are identified from high-definition T2-ZOOMit structural images, T2 images and SCS recruitment curve responses. Spinal segments are then reported in the T2 anatomical image acquired in each run. **Step 4**, Then motion correction is applied to each functional run in two stages. First, volumes are aligned to the averaged images using 3D rigid body realignment. Second, a slice-by-slice 2D realignment is applied to each volume. **Step 5**, The spinal cord (including gray and white matter) is automatically segmented from the motion corrected mean functional image, and then manually corrected. **Step 6**, Functional images from runs two and three are aligned to functional images of run one using the motion corrected mean images by landmarking the conus medullaris and any visible vertebra, using rigid transformations. **Step 7**, The motion corrected mean images are then coregistered to the PAM50 template using the conus medullaris landmark to extract the transformation matrices, using non-rigid transformations. **Step 8**, Physiological signals (heart rate and respiratory) acquired concomitantly to the fMRI volumes are used to model physiological noise (RETROICOR based procedure). Acquisition timings corresponding to motion corrected fMRI volume series and physiological noise regressors are submitted to a specific first level generalized linear model (GLM). Residuals from this GLM are then spatially smoothed, volume by volume with 3D gaussian kernel with full width at half maximum (FWHM) of 2×2×6mm^3^. Acquisition timings corresponding to the task-design and residuals resulting from noise removal are submitted to a specific first level GLM. A second level fixed effects analysis (subject level, task specific) is performed by combining the three runs. Activity pattern maps are uncorrected and thresholded (Z > 2.3, p < 0.01). **Step 9**, Transformations matrices found in step 7 are then applied to the resulting activity pattern maps of step 8 to arrive in PAM50 space. **Step 10**, Activity patterns from the three participants, are then compared and analyzed across scanning sessions using the spinal segments found in step 3.

**Extended data figure 9:**
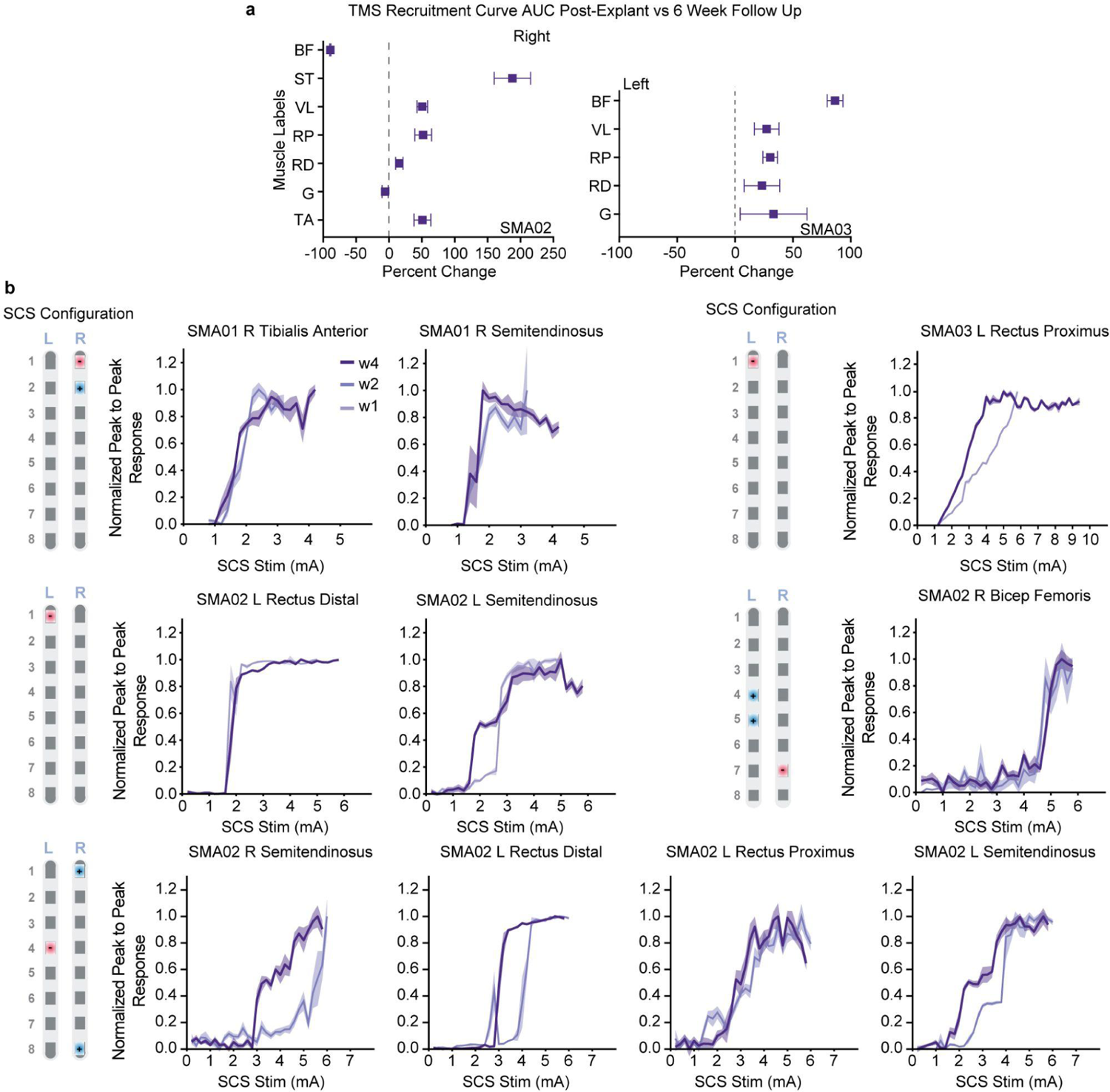
Assessment of spinal cord stimulation induced responses. **a**, Quantification of percent change in area under the curve of peak-to-peak TMS responses post-explant vs six week follow up across all intensities tested. Abbreviations: ST, Semitendinosus; BF, Bicep Femoris; VL, Vastus Lateralis; RP, Rectus Femoris Proximal; RD, Rectus Femoris Distal; G, Gastrocnemius; TA, Tibialis Anterior. **b**, The SCS parameters used and the resulting plots representing the normalized peak-to-peak response as a function of SCS amplitude. Plots are reporting the mean and 95% CI of the peak-to-peak responses. All error bars indicate the confidence interval, computed with bootstrap N=10,000 samples.

