## Supplementary information for "Targeted Stimulation of the Sensory Afferents Improves Motoneuron Function in Humans with Spinal Muscular Atrophy"

### Surgical Procedure

General anesthesia was induced using propofol and maintained using sevoflurane and propofol at levels that allowed for reliable somatosensory evoked potential monitoring. Short-acting paralytic was used for intubation, but no additional paralytic was given to facilitate intraoperative monitoring of SCS evoked EMG. All subjects were placed prone and affixed in a 3-pin Mayfield head holder. The back and neck were prepared and draped in typical sterile fashion and prophylactic antibiotics were administered. A small incision was made over the L2-L3 laminas using fluoroscopic guidance, and the tissue was dissected to expose the fascia. A Tuohy needle was inserted into the L2-L3 epidural interspace and used to guide the placement of a clinically approved 8 contact percutaneous spinal lead (PN 977A260, Medtronic). The two leads (left and right) were threaded rostrally and steered *in situ* using fluoroscopy towards the lateral aspect of the left and right spinal cord such that the most distal contact was positioned at the lateral aspect of the left and right spinal canal in the T11 vertebral body.

To confirm placement leads and ensure that we could recruit motor pools of the leg muscles, we delivered current controlled monopolar stimulation using an intraoperative neuromonitoring system (Xltek Protektor, Natus Medical). Stimulation pulses were delivered at 1-2 Hz on representative electrodes of the array and we measured compound muscle action potentials (CMAPs) using intramuscular needle electrodes (iliopsoas, gluteus, rectus femoris, vastus lateralis, semitendinous, tibialis anterior, and extensor hallucis longus ). We moved the leads in the rastro-caudal axis until we could selectively recruit the iliopsoas with the most rostral contact and the extensor hallucis longus with the most caudal contact. We also recorded contralateral activity to ensure that SCS did not induce cross-over effects between legs. Once satisfied with the lead placement, the Tuohy needle was removed, and the lead was sutured to the fascia to prevent lead migration.

To validate that stimulation was activating dorsal sensory afferent fibers and not directly recruiting ventral motor efferent fibers, we evaluated the stimulation frequency dependent response of CMAP amplitudes across several representative electrodes. Current amplitude was fixed at a level above the motor threshold (the amplitude above which CMAPs were reliably induced). Pulse frequency was then increased from 1-2 Hz up to 20 Hz and the relative, normalized, peak-to-peak amplitude of CMAP responses were compared.

To explant the arrays at the conclusion of the study period, the patients were prepared in a similar fashion to the implantation surgery. The upper thoracic incision was re-opened, and the lead wires were cut and removed proximally. The distal end of the leads were removed through the lateral exit wound and both incisions were closed.

### Stimulators

**Custom Stimulation Controller.** During the trial, SCS was delivered using a clinical grade, single channel, current controlled stimulator (DS8R, Digitimer) and a high-current compliant 1-to-8 multiplexer (D188, Digitimer) that we previously used. In previous a publication[1], we reported a more detailed description of the stimulator and the Matlab software used to control the stimulation. Here, we summarized the most important properties. Using the stimulator current could be delivered to any contact by connecting it to the multiplexer and selecting the associated output channel. A custom-built microcontroller-based (Arduino Due, Arduino) control unit sets pulse timing, amplitude, and output channel for each stimulus. Pulse width, inter-pulse interval, and waveform shape were fixed by the stimulator which ensured proper charge balancing and safe operation. Each pulse was a cathodic-first, biphasic square waveform with 400 us (SMA01), 500 us (SMA02) and 400 us (SMA03) monophase pulse width and 10 us inter-pulse interval. Cathodic and anodic phases were equivalent in amplitude and duration.

The control unit triggered each stimulus with a digital trigger pulse and set pulse amplitude using a continuous analog signal between 0-3.3 V. The DS8R hardware was configured for safety such that it could not produce amplitudes higher than 10.23 mA. Despite the stimulator comprising a single current source, the control unit's firmware enabled semi-synchronous stimulation across multiple channels by rapidly switching the output channel after each pulse. The time between pulses on separate channels was measured to be 2.2 ms, giving enough time for the multiplexer to fully switch output channels. During the study, we used this system to deliver stimulation on up to 4 separate spinal electrodes at up to 100 Hz. All programmable stimulation settings were configurable using a graphical user interface (GUI) developed in MATLAB which communicated with the control unit via a virtual serial port over a USB connection. Stimulation frequency, channel, duration, latency, and amplitude could all be configured manually via the GUI. Each channel could also be set to deliver a single pulse, a pulse train of fixed duration or pulse count, or continuous stimulation**.**

A custom command protocol was implemented to facilitate communication between the GUI and control unit. Communication was always initiated by the GUI with a command packet comprising the length in bytes of the packet, a 1-byte command, and 0-6 bytes of data. Possible commands included triggering or terminating stimulation, clearing the current configuration, reading or writing a parameter, configuring the microcontroller to accept new parameters (program mode), saving parameters, and an initialization handshake. When writing parameters, the length and command bytes were followed by the parameter to be set, the channel (if applicable), and the value to be written. When reading parameters, the data payload comprised only the parameter to be read. All commands were followed by a response packet from the microcontroller comprising the length of the packet, an echo of the command received, a data payload if applicable (for example when reading parameters), and a status byte indicating whether the command was executed correctly.

**Wireless stimulator.** During the trial SCS was also delivered using the Medtronic Model 97725 Wireless External Neurostimulator with Bluetooth wireless technology. It is a multi-programmable device that delivers stimulation through 1 or more leads. The stimulation settings are stored in programs where specific combinations of pulse width, rate and intensity settings are configured for specific electrode combinations (up to 16 electrodes per program), with up to 4 programs being running simultaneously. When stimulating with more than one program pulses are delivered sequentially, first a pulse from one program and then a pulse from the next program. Pulse width, intensity, cycling, and electrode polarity for each program within a group can have different values. Rate, rate limits, pulse width limits, and intensity limits for each program within a group have the same values. Programs were controlled from an external touchscreen Samsung tablet with Intellis™ clinician programmer app A710.

**EMG Acquisition**

To assess muscle activity during movement, surface electromyography (sEMG) was recorded using a wireless EMG system (Trigno, Delsys Inc.). Up to 14 synchronized wireless sensors (Avanti Trigno, Delsys Inc.) were used to amplify, digitize, and wirelessly transmit EMG signals to a base station unit. Each sensor sampled the analog signal at 1925.925 Hz and applied a hardware bandpass filter of 20-800 Hz**.** Once the signals were received by the base station, they were converted back to an analog waveform and resampled at 8000 Hz by a data acquisition system (PCI-6255, National Instruments) for synchronization with other task events. The Trigno system has a known, fixed wireless latency of 59.6 ms.

At the beginning of each experimental session, the legs were cleaned using mildly abrasive skin preparation gel and isopropyl alcohol. Skin safe adhesive was used to secure the EMG sensors to the subject's arm. Depending on the muscles of interest for a particular experiment, we recorded from up to 14 individual muscles (7 each leg) of the legs; including the rectus femoris (proximal and distal), vastus lateralis, bicep femoris, semitendinosus, gastrocnemius, and tibialis anterior whose locations were identified by palpation according to the SENIAM's recommendations[2]. The rectus femoris proximal location was used as a proxy for the iliopsoas (hip flexor) muscle as this can not be recorded by a surface EMG sensor. Sensors were then carefully removed at the end of each session. Placement of EMG sensors were marked after removal to assure accurate and repeatable measurements across sessions.

**Figure Generation**

All figures were composed in Adobe Illustrator CC v26.0-26.3.1.

**Supplementary Table 1**: List of all primary and secondary outcomes of the clinical study

| **Primary Objectives:** | **Measure** | **Success Criteria** | **Timeframe** |
|---|---|---|---|
| Muscle weakness – isometric torque | Isometric torque will be measured produced by the hip during hip flexion | At least 20% increased torque production over SCS-on vs SCS-off baseline as measured during single-joint isometric torque. | During the 4 week implant period |
| Safety | Record all adverse events | No serious adverse events related to the | Until 2 weeks post-explant |

| | | | |
|---|---|---|---|
| | | use of electrical stimulation reported | |
| **Secondary Objectives:** | **Measure** | **Success Criteria** | **Timeframe** |
| Muscle weakness | Surface EMGs produced by subjects during isometric movements of the hip, knee, and ankle | At least 20% increase in EMG RMS in SCS-on vs SCS-off | During the 4 week implant period |
| Motor function – Range of Motion | Range of motion using the HUMAC Norm at the hip and knee joints | At least 20% increase in ROM in SCS-on vs SCS-off | During the 4 week implant period |
| Motor function – 6 Minute Walk Test | Distance walked | At least 24m increased distance traveled between SCS-off and SCS-on | During the 4 week implant period |
| Motor function – Revised Hammersmith Score | Scoring on the standardized test | At least 2 point improvement in scores between SCS-on and SCS-off | During the 4 week implant period |
| Motor Function – Motor Function Measure 32 | Scoring on the standardized test | At least 2 point improvement in scores between SCS-on and SCS-off | During the 4 week implant period |
| Discomfort/pain | Participants are asked to report perceived discomfort, utilizing a 1-10 scale. Lower numerical values are equated with lower perceived discomfort. | Participants do not report discomfort or pain at stimulation amplitudes that are required to obtain motor responses. | During the 4 week implant period |
| Clinical Global Impression Scale | Feedback on how the technology is working and what the participants will improve. | | During the 4 week implant period |

**Structural MRI of the Lumbosacral Spine**

All participants underwent structural MRI of the lumbosacral spine on a 3T MRI scanner

(Magnetom Prisma$^{fit}$, Siemens Healthineers) with a 32-channel spine array coil at the BRIDGE center (Carnegie Mellon University - CMU-Pitt BRIDGE Center, RRID:SCR_023356). Participants were placed into the MRI scanner in a supine position with a pad underneath their knees to eliminate the curve of the lumbar spine and arms at their sides. The standard MRI protocol comprised the following two pulse sequences, all used without gadolinium-based contrast agent administration: a) high-resolution sagittal T2-weighted SPACE sequence (single slab 3D turbo spin echo sequence with a slab selective, variable excitation pulse, repetition time (TR), 1500 ms; echo time (TE), 135 ms; voxel size, 0.4×0.4×0.8 $mm^3$); b) 2D transverse T2-weighted SPACE with ZOOMit (dynamic excitation pulses to achieve selective/zoomed field-of-view) software (TR, 3080 ms; TE, 106 ms; voxel size, 0.9×0.9×0.5 $mm^3$). Structural scanning took less than 16 minutes in total. Detailed acquisition parameters for the two MRI pulse sequences used to image the medullary cone and nerve roots of the participants' lumbar spine are in **Supplementary Table 2**. Prior to each pulse sequence acquisition, manual shim boxes were optimized to correct for magnetic field inhomogeneities.

**Supplementary Table 2**: MRI Sequence Parameters

| **Sequence Parameters** | **T2-weighted SPACE** | **T2-weighted SPACE with ZOOMit** |
|---|---|---|
| Plane | Sagittal | Transverse |
| Number of Slices | 64 | 160 |
| Slice Thickness (mm) | 0.80 | 0.5 |
| Field of View (mm) | 260 x 260 | 300 x 300 |
| Acquisition Matrix | 320 x 301 | 320 x 115 |
| Voxel Size (mm) | 0.4 x 0.4 x 0.8 | 0.9 x 0.9 x 0.5 |
| K-Space Sampling | GRAPPA | Spiral |
| Phase-Encoding Direction | Head to Feet | Anterior to Posterior |
| Repetition Time (ms) | 1500 | 3080 |
| Echo time (ms) | 135 | 106 |
| Turbo Factor | 88 | 270 |
| Number of Signal Averages | 1.4 | 1.4 |
| Flip Angle (degrees) | 140 | 100 |
| Bandwidth (Hz/pixel) | 625 | 579 |
| Acquisition Tim (min:sec) | 5:54 | 9:16 |

**Spinal cord fMRI**

We performed lumbosacral spinal cord functional magnetic imaging (fMRI) to obtain visualization of the motor neurons innervating specific muscles. Motor neurons control the movement of muscle which were recruited through active extension and flexion of the knee joint. The fMRI pre-processing, processing and analysis pipeline were based on recent cervical spinal cord fMRI studies and lumbar spinal cord studies[3–8]. We tailored this pipeline to image the lumbar spinal cord.

**fMRI data acquisition**

Participants were already installed in the MRI scanner for structural imaging and position did not change. The participants were instructed to relax, remain still and to breathe normally. SMA01 underwent three scanning sessions, while SMA02 and SMA03 underwent four scanning sessions, involving active mobilization of the knee joint. Functional acquisitions were performed using a gradient-echo echo-planar sequence with a ZOOMit field-of-view imaging, with repetition time (TR), 2500 ms; echo time (TE), 34 ms; field of view (FOV), 48x144 mm; flip angle, 80°; voxel size, 1.0 x 1.0 x 3.0mm$^3$; and 32 axial slices per volume. The second to last bottom slice was placed at the tip of the conus medullaris. Manual shimming adjustments focused on the spinal cord were done to adjust field homogeneity. Physiological data (respiratory and cardiac signals) were directly acquired using MRI compatible photoplethysmography and respiratory belt (Biopac MP150 system, California, USA).

**Supplementary Table 3**: Adverse events related to the study

| Participant ID | Number and type of adverse events | Serious |
|---|---|---|
| SMA01 | N=2; Fall, resolved, no sequalae | No |
| SMA02 | N=2; Falls, resolved, no sequalae | No |
| SMA03 | N=1; Fall, resolved, no sequalae | No |
